# Systematic review of imaging tests to predict the development of rheumatoid arthritis in people with unclassified arthritis

**DOI:** 10.1101/2021.03.16.21253692

**Authors:** P de Pablo, J Dinnes, S Berhane, A Osman, Z Lim, A Coombe, K Raza, A Filer, JJ Deeks

**Author notes:** J Dinnes and P de Pablo contributed equally to this work. Correspondence to: J Dinnes, Institute of Applied Health Research, University of Birmingham, Edgbaston, Birmingham, UK B15 2TT.

## Abstract

**Objectives:** To estimate and compare the diagnostic accuracy of magnetic resonance imaging (MRI) and ultrasound, for the prediction of rheumatoid arthritis (RA) in unclassified arthritis (UA).

**Methods:** MEDLINE, Embase and BIOSIS were searched from 1987 to May 2019. Studies evaluating any imaging test in participants with UA were eligible. Reference standards were RA classification criteria or methotrexate initiation. Two authors independently extracted data and assessed validity using QUADAS-2. Sensitivities and specificities were calculated for each imaging characteristic and joint area. Summary estimates with 95% confidence intervals (CI) were estimated where possible.

**Results:** Nineteen studies were included; 13 evaluated MRI (n=1,143; 454 with RA) and 6 evaluated ultrasound (n=531; 205 with RA). Studies were limited by unclear recruitment procedures, inclusion of patients with RA at baseline, differential verification, lack of blinding and consensus grading. Study heterogeneity largely precluded meta-analysis, however summary sensitivity and specificity for MRI synovitis in at least one joint were 93% (95% CI 88%, 96%) and 25% (95% CI 13%, 41%) (3 studies). Specificities may be higher for other MRI characteristics but data are limited. Ultrasound results were difficult to synthesise due to different diagnostic thresholds and reference standards.

**Conclusions:** The evidence for MRI or ultrasound as single tests for predicting RA in people with UA is heterogeneous and of variable methodological quality. Larger studies using consensus grading and consistently defined RA diagnosis are needed to identify whether combinations of imaging characteristics, either alone or in combination with other clinical findings, can better predict RA in this population.

**Systematic review registration:** PROSPERO CRD42020158239.

**Key messages:**

- To date, the diagnostic accuracy of imaging tests for the earlier identification of RA has not been systematically assessed. We conducted a systematic review to estimate, and if possible compare, the accuracy of MRI and ultrasound for predicting the diagnosis of rheumatoid arthritis in people with unclassified arthritis.
- In this systematic review of 13 studies of MRI (1,143 participants) and 6 studies of ultrasound (531 participants), study quality was highly variable with considerable variation in populations, diagnostic thresholds and reference standards limiting potential for meta-analysis.
- Individual MRI imaging characteristics demonstrated either high sensitivity (with low specificity) or high specificity (with low sensitivity) with inconsistent results between studies. Similar heterogeneity in results was observed for ultrasound but with considerably fewer data.
- Imaging can identify subclinical inflammatory changes in joint areas where no synovitis is apparent, which may be useful in identifying the aetiology of symptoms. However, larger studies using consistent scoring systems for imaging interpretation and definition of RA are needed to identify the extent to which imaging findings alone can predict the development of RA. Until then, imaging should be interpreted in light of other findings.

## INTRODUCTION

Despite major treatment advances, rheumatoid arthritis (RA) is associated with long-term morbidity and accelerated mortality^1^. Uncontrolled RA also impacts on daily activities and quality of life^2 3^ and can confer a substantial socioeconomic burden and loss in productivity^4–6^. Early diagnosis and treatment of RA can prevent joint damage, and ensure that people remain productive and in work^7^. Evidence for a therapeutic window of opportunity early in the course of the disease is accumulating^8 9^, and has focused attention on earlier identification of which patients will go on to develop RA ^10^. However, among those who present with unclassified arthritis (UA), about 60% have a self-limiting disease while 40% develop a chronic persistent arthritis (of which only a proportion is RA)^11^, making the accurate and timely diagnosis of RA in patients with UA an important aim.

There is no single test to confirm the diagnosis of RA. Classification criteria developed by the American College of Rheumatology (ACR 1987) (most recently in collaboration with the European League Against Rheumatism (ACR/EULAR 2010))^12 13^ are often used as a proxy for diagnosis. The classification criteria have historically relied on clinical signs and symptoms, rheumatoid factor (RF) and radiographic changes^13^, with the 2010 revision incorporating inflammation markers (ESR and CRP) and anti-cyclic citrullinated antibodies (anti-CCP)^12^. Because of enhanced sensitivity to detect early disease, the ACR/EULAR 2010 criteria may lead to early and appropriate initiation of anti-rheumatic therapy among some individuals, but at the cost of unnecessary treatment in others^14^. The development of more accurate diagnostic strategies for RA in patients with very early joint symptoms is therefore of critical importance.

A number of clinical prediction models using various clinical and serological criteria have been developed^15–17^, however an array of imaging modalities with the potential to improve early diagnosis are also available^18 19^. Musculoskeletal ultrasound and magnetic resonance imaging (MRI) can identify joint inflammation and structural changes even in those with normal radiographs^20–23^ and are increasingly used in clinical practice^24^ or for research purposes ^25^. Positron emission tomography (PET)^26 27^ and single-photon emission computed tomography (SPECT)^28^ are also being investigated in RA, along with novel approaches such as fluorescence optical imaging^29–32^ and optical spectral transmission imaging^33 34^.

The diagnostic accuracy of imaging tests for the earlier identification of RA has not yet been systematically assessed. We conducted a systematic review to estimate, and if possible compare, the accuracy of imaging tests in the prediction of RA in newly presenting patients with UA.

## METHODS

We followed published methods for systematic reviews^35^ and report our findings according to the Preferred Reporting Items for Systematic Reviews and Meta-Analyses (PRISMA) extension for diagnostic test accuracy studies^36^.

### Data sources

We searched MEDLINE, Embase and BIOSIS from 1987 to 2^nd^ May 2019. Full search strategies are available (supplementary appendix 1). No language restrictions were applied. Reference lists of systematic reviews and included study reports were also screened.

### Study selection

Study selection was undertaken independently by two reviewers; disagreements were resolved by discussion. Studies evaluating any imaging test characteristic in participants with UA with at least one clinically swollen, or with at least one clinically swollen or tender joint, were eligible for inclusion if they provided a cross-tabulation of imaging results at baseline against the subsequent diagnosis of RA (defined as fulfilment of ACR 1987^13^ or ACR/EULAR 2010^12^ classification criteria or by the initiation of methotrexate or other DMARD treatment), at least three months later. Studies in populations with clinically suspect arthralgia^37^ or where UA was defined by joint tenderness, or including more than 50% of participants with RA at baseline were excluded unless subgroup data for UA were presented. Case-control studies, conference abstracts and studies recruiting less than five participants with or without a final diagnosis of RA were also excluded. A list of excluded studies with reasons for exclusion is available on request. Authors of eligible studies were contacted when they presented insufficient data to allow for the construction of 2 × 2 contingency tables.

### Data collection, quality assessment and analysis

Two reviewers independently extracted data using a pre-specified and piloted data extraction form and assessed study quality using the Quality Assessment of Diagnostic Accuracy Studies-2 tool (QUADAS-2) ^38^ (supplementary appendix 2). Any disagreements were resolved by consensus. Each study should ideally prospectively recruit a representative sample of participants with UA and should exclude those who meet RA classification criteria at baseline. Blinding of imaging test interpretation and of final diagnosis of RA should be implemented and standard scoring systems such as RAMRIS for MRI^39 40^, recent EULAR-OMERACT^41–44^ or previous OMERACT definitions^42 45–48^ for grading ultrasound synovitis in RA and/or widely used consensus definitions for ultrasound^19 41 49–51^ should be used.

Estimates of sensitivity and specificity from each study were plotted on coupled forest plots for each imaging characteristic and joint area imaged. Where two or more studies used the same scoring system and reference standard, summary sensitivities and specificities with 95% confidence intervals (95% CI) were obtained using the bivariate hierarchical model^52^. Due to paucity of studies, the models were simplified by assuming no correlation between sensitivity and specificity estimates and by setting near-zero variance estimates of the random effects to zero^53^. Study heterogeneity was examined by inspection of forest plots; no formal investigation was conducted due to data scarcity.

Plots were produced using RevMan 5.3 (Nordic Cochrane Centre) and analyses undertaken with STATA 16 software using meqrlogit (bivariate hierarchical models), or blogit commands (fixed effect logistic regression). Absolute differences in sensitivities or specificities were derived using nlcom.

## RESULTS

A total of 204 records were selected for full-text assessment from 10,812 unique references (Figure 1). Corresponding authors of 26 publications (13 conference abstracts and 13 full text papers) were contacted; information was supplied by 16, resulting in inclusion of three^56 57 70^ and exclusion of 12 ^21 73–84^. Common reasons for exclusion were ineligible study participants, ≥50% with RA; 74/185, 40%) or lack of follow up (35/185, 19%). Other exclusion reasons are displayed on Figure 1 and detailed in supplementary appendix 2.

**Figure 1.**
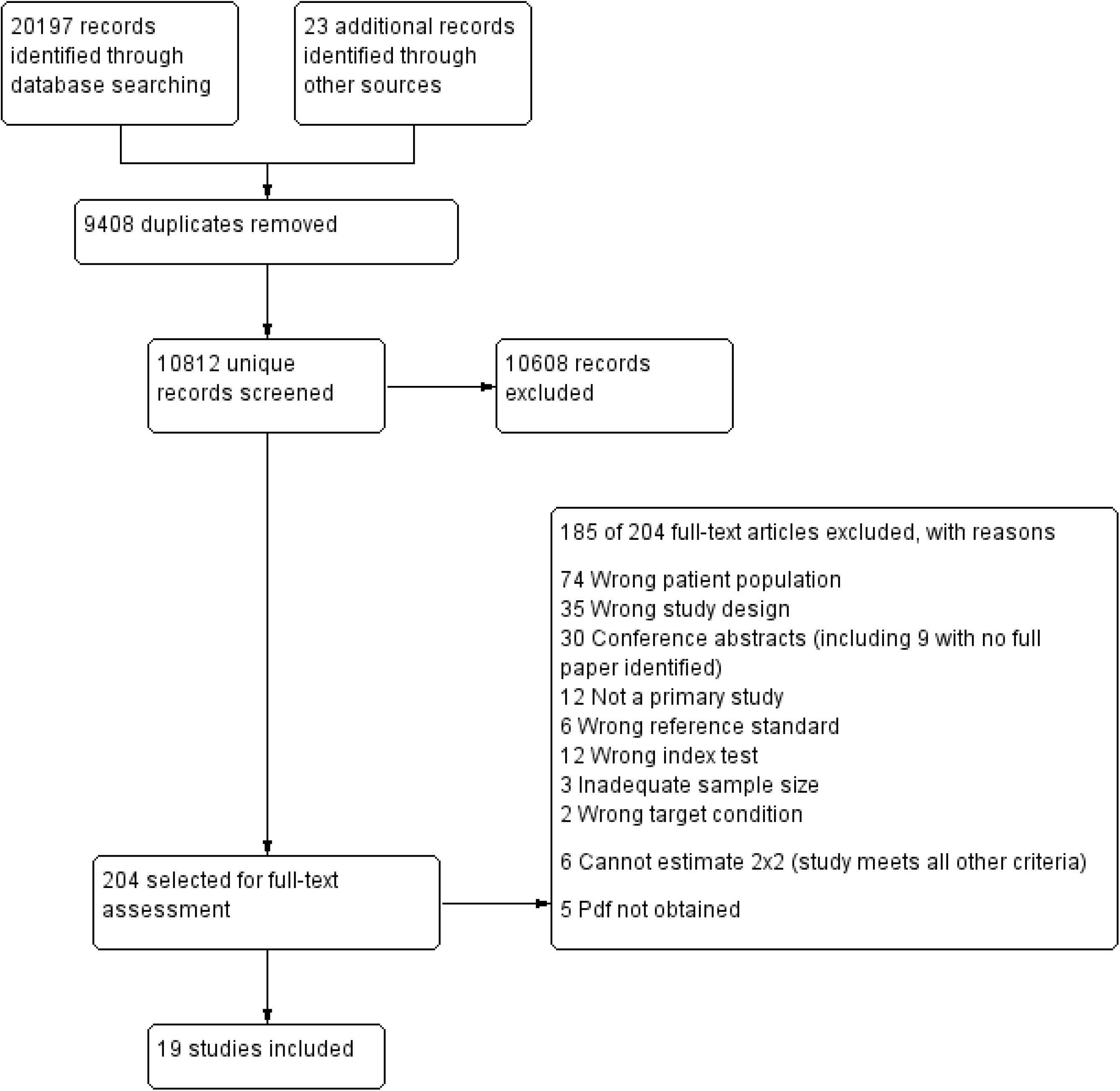
PRISMA Flow diagram

Nineteen studies (19/204, 9%) met inclusion criteria. Table 1 provides a summary of key study characteristics and further imaging details are shown on Table 3. Thirteen studies (N=1,143, RA n=454) evaluated the accuracy of individual MRI characteristics ^55 59–63 65–71^, and six studies (N=531, RA n=205) evaluated ultrasound^54 56–58 64 72^. One study reported data for scintigraphy^60^. Median sample size was 81 (inter-quartile range (IQR) 39, 119) and median prevalence of RA at the end of study follow up was 38% (29%, 51%), with 45% (29%, 58%) for MRI and 34% (25, 40%) for ultrasound.

**Table 1.**
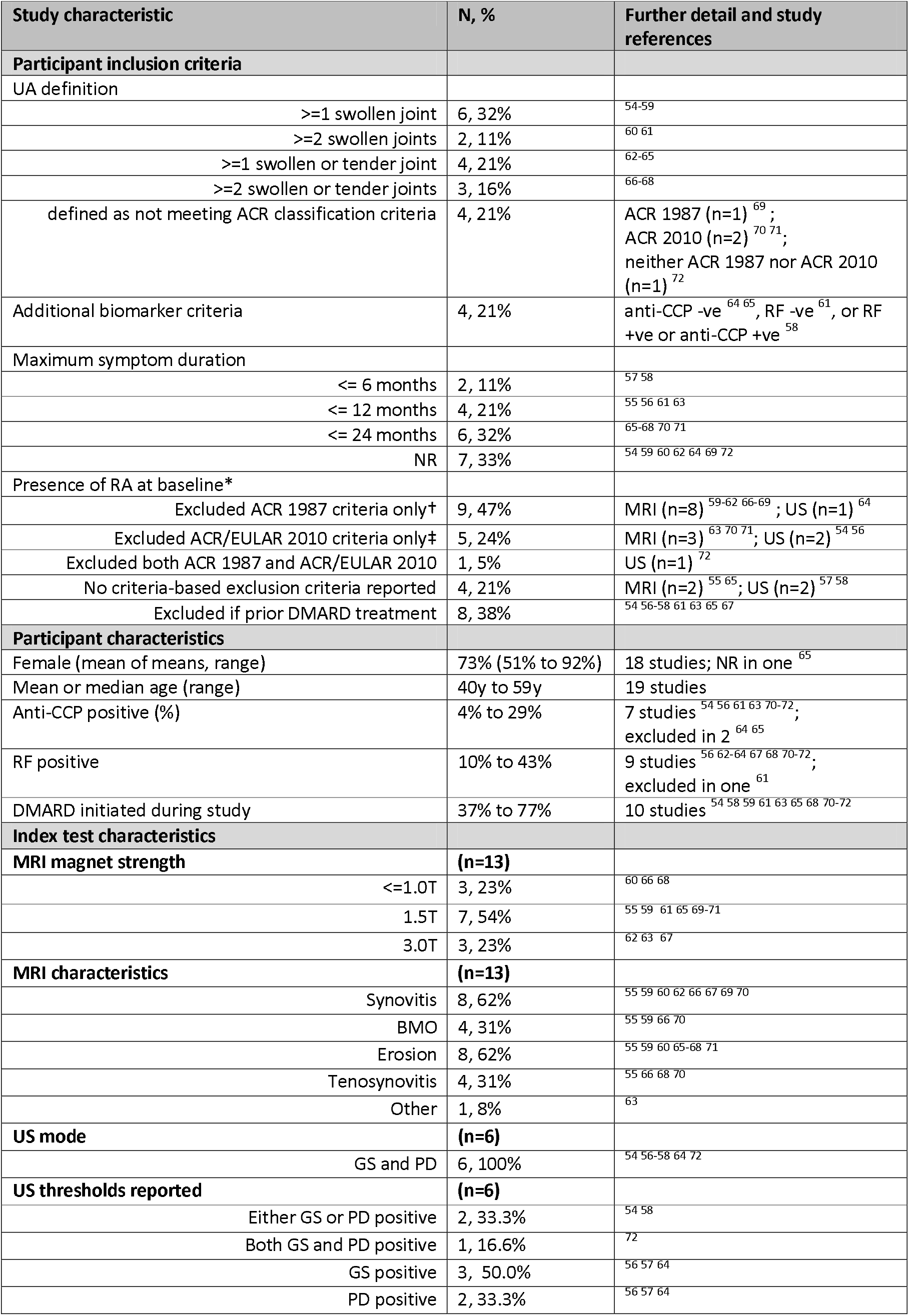
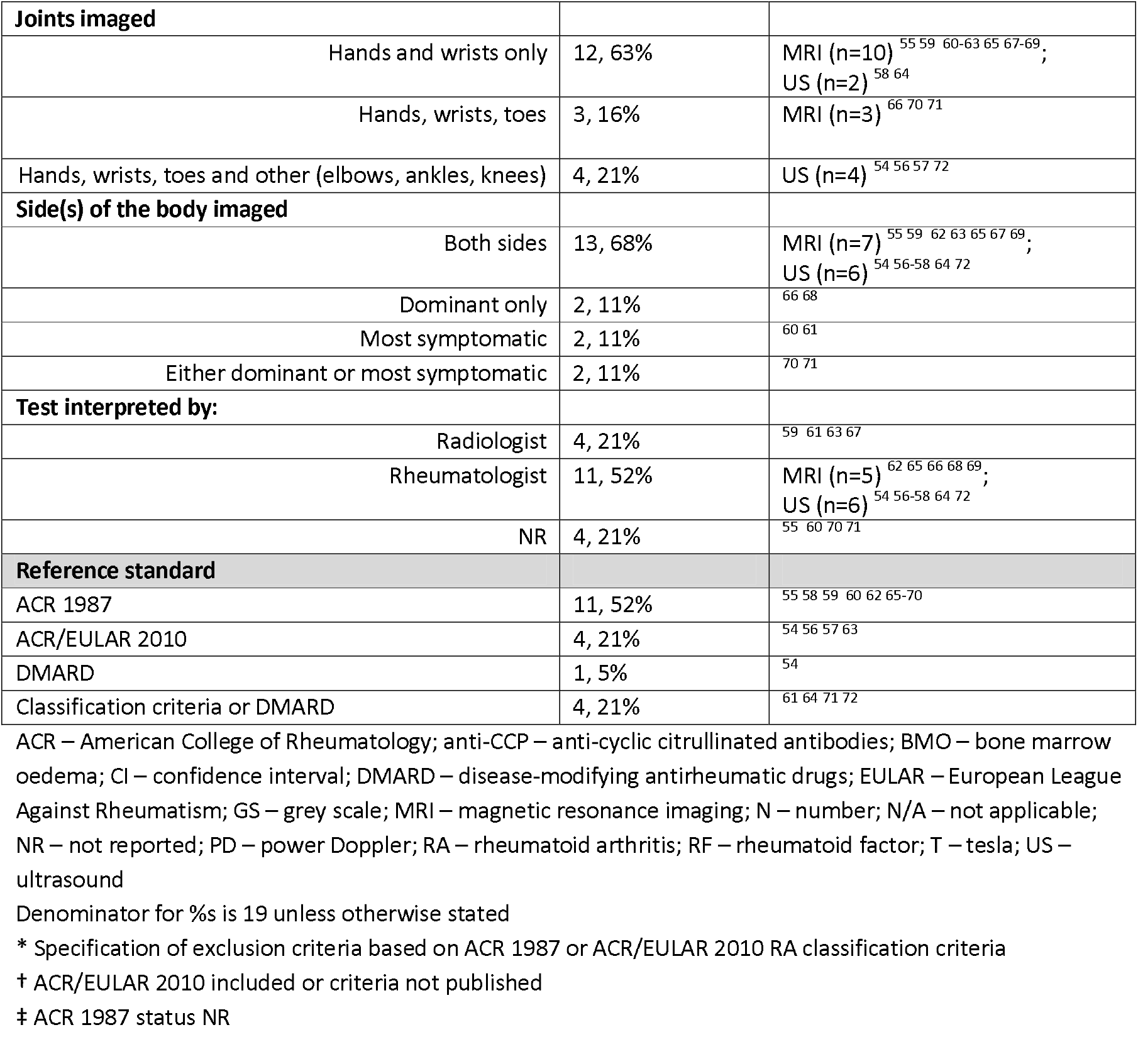
Summary characteristics across studies

| Study characteristic | N, % | Further detail and study references |
| --- | --- | --- |
| **Participant inclusion criteria** |  |  |
| UA definition |  |  |
| >=1 swollen joint | 6, 32% | 54-59 |
| >=2 swollen joints | 2, 11% | 60 61 |
| >=1 swollen or tender joint | 4, 21% | 62-65 |
| >=2 swollen or tender joints | 3, 16% | 66-68 |
| defined as not meeting ACR classification criteria | 4, 21% | ACR 1987 (n=1)<sup>69</sup>; ACR 2010 (n=2)<sup>70 71</sup>; neither ACR 1987 nor ACR 2010 (n=1)<sup>72</sup> |
| Additional biomarker criteria | 4, 21% | anti-CCP -ve<sup>64 65</sup>, RF -ve<sup>61</sup>, or RF +ve or anti-CCP +ve<sup>58</sup> |
| Maximum symptom duration |  |  |
| <= 6 months | 2, 11% | 57 58 |
| <= 12 months | 4, 21% | 55 56 61 63 |
| <= 24 months | 6, 32% | 65-68 70 71 |
| NR | 7, 33% | 54 59 60 62 64 69 72 |
| Presence of RA at baseline* |  |  |
| Excluded ACR 1987 criteria only† | 9, 47% | MRI (n=8)<sup>59-62 66-69</sup>; US (n=1)<sup>64</sup> |
| Excluded ACR/EULAR 2010 criteria only‡ | 5, 24% | MRI (n=3)<sup>63 70 71</sup>; US (n=2)<sup>54 56</sup> |
| Excluded both ACR 1987 and ACR/EULAR 2010 | 1, 5% | US (n=1)<sup>72</sup> |
| No criteria-based exclusion criteria reported | 4, 21% | MRI (n=2)<sup>55 65</sup>; US (n=2)<sup>57 58</sup> |
| Excluded if prior DMARD treatment | 8, 38% | 54 56-58 61 63 65 67 |
| **Participant characteristics** |  |  |
| Female (mean of means, range) | 73% (51% to 92%) | 18 studies; NR in one<sup>65</sup> |
| Mean or median age (range) | 40y to 59y | 19 studies |
| Anti-CCP positive (%) | 4% to 29% | 7 studies<sup>54 56 61 63 70-72</sup>; excluded in 2<sup>64 65</sup> |
| RF positive | 10% to 43% | 9 studies<sup>56 62-64 67 68 70-72</sup>; excluded in one<sup>61</sup> |
| DMARD initiated during study | 37% to 77% | 10 studies<sup>54 58 59 61 63 65 68 70-72</sup> |
| **Index test characteristics** |  |  |
| MRI magnet strength | (n=13) |  |
| <=1.0T | 3, 23% | 60 66 68 |
| 1.5T | 7, 54% | 55 59 61 65 69-71 |
| 3.0T | 3, 23% | 62 63 67 |
| MRI characteristics | (n=13) |  |
| Synovitis | 8, 62% | 55 59 60 62 66 67 69 70 |
| BMO | 4, 31% | 55 59 66 70 |
| Erosion | 8, 62% | 55 59 60 65-68 71 |
| Tenosynovitis | 4, 31% | 55 66 68 70 |
| Other | 1, 8% | 63 |
| US mode | (n=6) |  |
| GS and PD | 6, 100% | 54 56-58 64 72 |
| US thresholds reported | (n=6) |  |
| Either GS or PD positive | 2, 33.3% | 54 58 |
| Both GS and PD positive | 1, 16.6% | 72 |
| GS positive | 3, 50.0% | 56 57 64 |
| PD positive | 2, 33.3% | 56 57 64 |
| **Joints imaged** |  |  |
| Hands and wrists only | 12, 63% | MRI (n=10)<sup>55 59 60-63 65 67-69</sup>;<br>US (n=2)<sup>58 64</sup> |
| Hands, wrists, toes | 3, 16% | MRI (n=3)<sup>66 70 71</sup> |
| Hands, wrists, toes and other (elbows, ankles, knees) | 4, 21% | US (n=4)<sup>54 56 57 72</sup> |
| **Side(s) of the body imaged** |  |  |
| Both sides | 13, 68% | MRI (n=7)<sup>55 59 62 63 65 67 69</sup>;<br>US (n=6)<sup>54 56-58 64 72</sup> |
| Dominant only | 2, 11% | <sup>66 68</sup> |
| Most symptomatic | 2, 11% | <sup>60 61</sup> |
| Either dominant or most symptomatic | 2, 11% | <sup>70 71</sup> |
| **Test interpreted by:** |  |  |
| Radiologist | 4, 21% | <sup>59 61 63 67</sup> |
| Rheumatologist | 11, 52% | MRI (n=5)<sup>62 65 66 68 69</sup>;<br>US (n=6)<sup>54 56-58 64 72</sup> |
| NR | 4, 21% | <sup>55 60 70 71</sup> |
| **Reference standard** |  |  |
| ACR 1987 | 11, 52% | <sup>55 58 59 60 62 65-70</sup> |
| ACR/EULAR 2010 | 4, 21% | <sup>54 56 57 63</sup> |
| DMARD | 1, 5% | <sup>54</sup> |
| Classification criteria or DMARD | 4, 21% | <sup>61 64 71 72</sup> |

ACR – American College of Rheumatology; anti-CCP – anti-cyclic citrullinated antibodies; BMO – bone marrow oedema; CI – confidence interval; DMARD – disease-modifying antirheumatic drugs; EULAR – European League Against Rheumatism; GS – grey scale; MRI – magnetic resonance imaging; N – number; N/A – not applicable; NR – not reported; PD – power Doppler; RA – rheumatoid arthritis; RF – rheumatoid factor; T – tesla; US – ultrasound
Denominator for %s is 19 unless otherwise stated
\* Specification of exclusion criteria based on ACR 1987 or ACR/EULAR 2010 RA classification criteria
† ACR/EULAR 2010 included or criteria not published
‡ ACR 1987 status NR

UA was defined as clinical synovitis in at least one joint (6/19, 32%) ^54–59^, more than one swollen joint (2, 11%) ^60 61^ and at least one (4, 21%)^62–65^, or more than one (3, 16%)^66–68^ swollen or tender joint of the hand or wrist. Four studies (21%) only stipulated that participants not meet RA classification criteria at baseline with no further detail^69–71 72^. Four studies (21%) restricted inclusion to anti-CCP negative^64 65^, RF negative^61^, or either RF or anti-CCP positive^58^ participants. Fifteen studies (79%) excluded participants meeting one set of RA classification criteria at baseline (n=9 ACR 1987^59–62 64 66–69^; n=5 ACR/EULAR 2010^54 56 63 70 71^), or either set of criteria (Table 1).^72^

MRI magnet strength ranged between 0.2T^60^ −3.0T^62 63 67^, 7 studies used 1.5T^55 59 61 65 69–71^. MRI synovitis (8/13, 62%) and erosion (8/13, 62%) were commonly assessed. 69% (9/13) studies used RAMRIS^59 61 63 65–67 71 85 86^.

Four studies (4/6, 67%)^54 57 58 64 72^ assessed ultrasound tenosynovitis (TS) defined as absent or present^19 48 87^ or based on a consensus-based US score for TS in RA^45 47^. All ultrasound evaluations included both grey scale (GS) synovitis and power dower (PD) activity. Synovitis was scored using similar semiquantitative grading scales^19 49-51^. Two studies^54 58^ used the OMERACT definition of synovitis^45 46^ and none used the more recent EULAR-OMERACT combined scoring system for grading synovitis in RA^41–44^. There was considerable variation between studies in joint sets imaged and use of bilateral imaging (Table 1).

### Validity and applicability of the evidence (QUADAS-2)

Six studies were rated low risk of bias for participant selection, ^54 55 61 62 71 72^ and two^57 65^ as high risk (Figure 2). Risk of bias could not be judged in 11 studies due to unclear recruitment procedures^56 58–60 63 64 66–69^ or possible inclusion of participants meeting reference standard criteria at baseline^58 70^. Study eligibility criteria did not match our UA definition in 10 studies^58 60–68^, including three reporting median symptom duration over 12 months^60 64 67^ leading to concerns about applicability of the study population. The generalisability of the study population could not be judged in four studies^69–72^.

**Figure 2.**
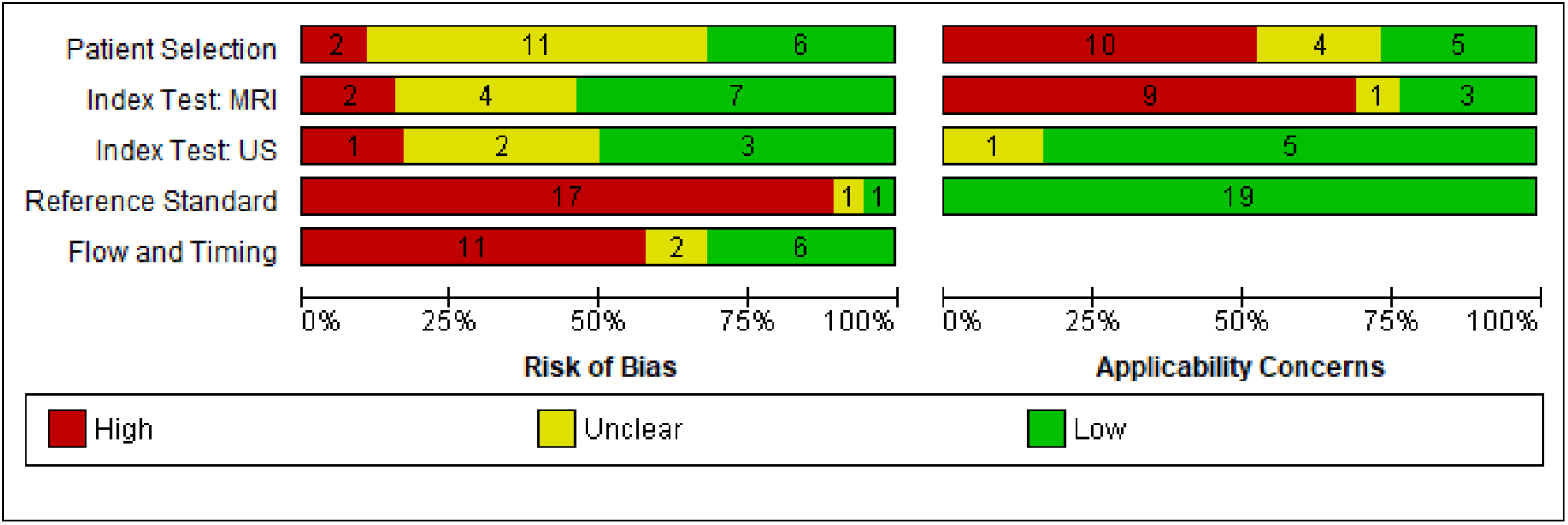
Risk of bias and applicability concerns graph

Risk of bias for the index test was low in 53% (10/19) of studies ^55 57 58 61 62 64 65 69–71^. Risk was high for two MRI evaluations due to lack of pre-specified diagnostic thresholds^63 67^ and one of ultrasound due to lack of blinding^72^. Blinding to baseline clinical findings was not clearly reported in two ultrasound^54 56^ and four MRI evaluations^59 60 66 88^. Ultrasound (5/6) was interpreted by a single experienced or trained sonographer or rheumatologist, using clearly defined criteria for interpretation ^54 56–58 64 72^. We had high (9/13) ^59 60 62 65 67–71^, or unclear (1/13)^55^ concerns for the applicability of MRI evaluations; 7 studies reporting consensus or mean results^59 62 65 67 69–71^ and three provided no MRI scoring details^60 68 69^. Seven studies did not describe MRI interpreter experience^55 60 62 65 68 69 71^.

Reference standard assessment was explicitly blinded to imaging test results in three studies ^64 67 73^ (one scored unclear risk of bias overall because of a combined reference standard^64^). Lack of blinding was reported (n=1)^55^ or was inferred in 16 studies ^54 56–63 65 66 68–72^.

There was high risk of bias for participant flow in 58% of studies (11/19), due to exclusion of participants from analysis ^54–56 62 65 66 68 69^ or differential verification ^61 71 72^. No unevaluable images were reported.

### Study synthesis

Sensitivities and specificities are presented by reference standard and imaging characteristic (Figures 3–5; supplementary Figures 2-4).

**Figure 3.**
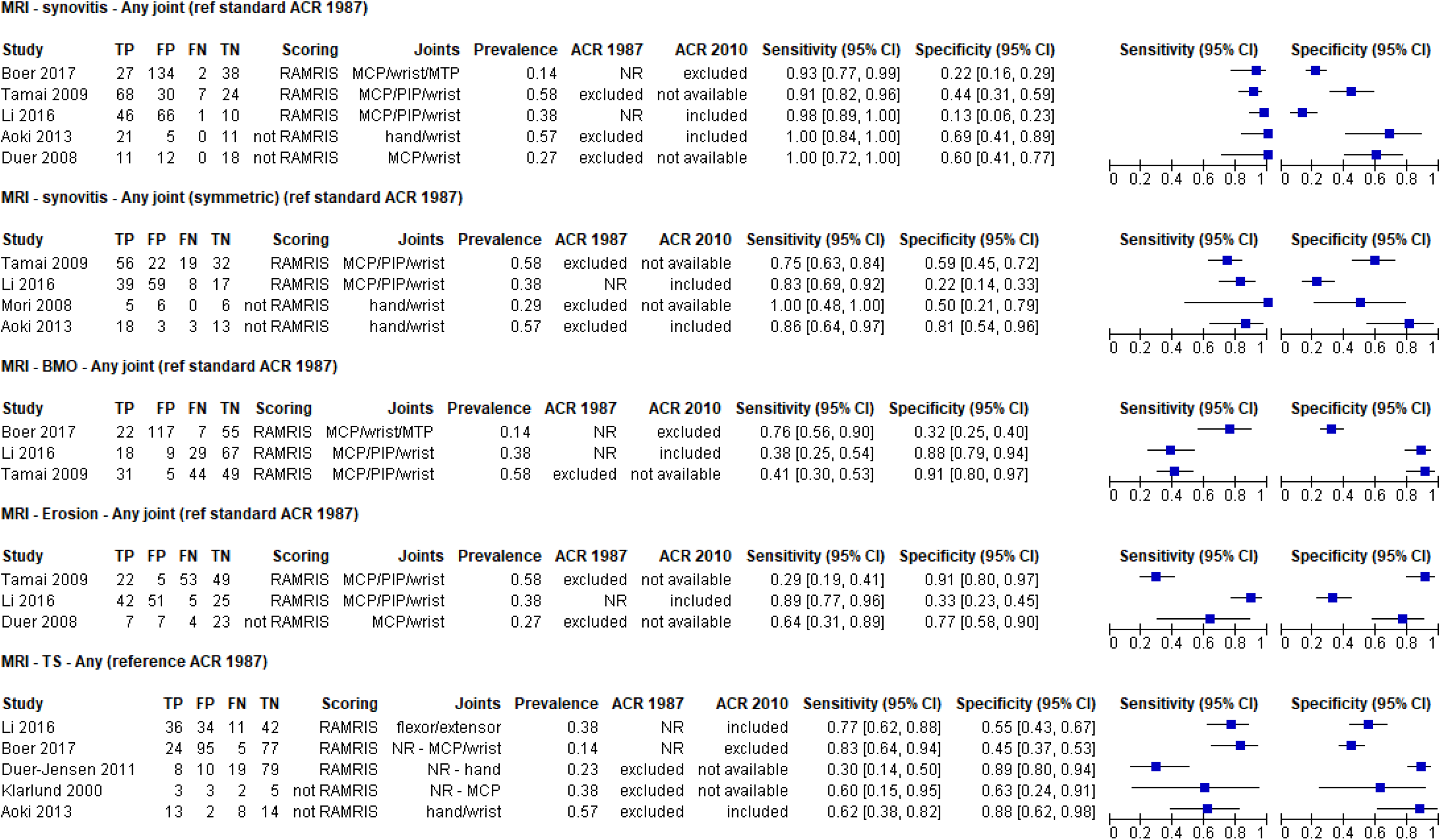
Forest plot of sensitivities and specificities from studies evaluating individual MRI characteristics (any joint positive against ACR 1987 reference standard)

#### MRI performance

Eight studies ^55 59 60 62 66 68–70^ evaluated MRI in any joint using ACR 1987 criteria as the reference standard (Figure 3). Four studies^55 59 66 70^ used RAMRIS and were eligible for statistical pooling.

Summary sensitivity and specificity for synovitis was 93% (95% CI 88% to 96%) and 25% (95% CI 13% to 41%) (n=3^55 59 70^; 453 participants [151 RA]) and for symmetric synovitis 78% (95% CI 70% to 84%) and 39% (95% CI 17% to 67%), respectively (n=2^55 59^; 252 [122]) (Table 2).

**Table 2.**
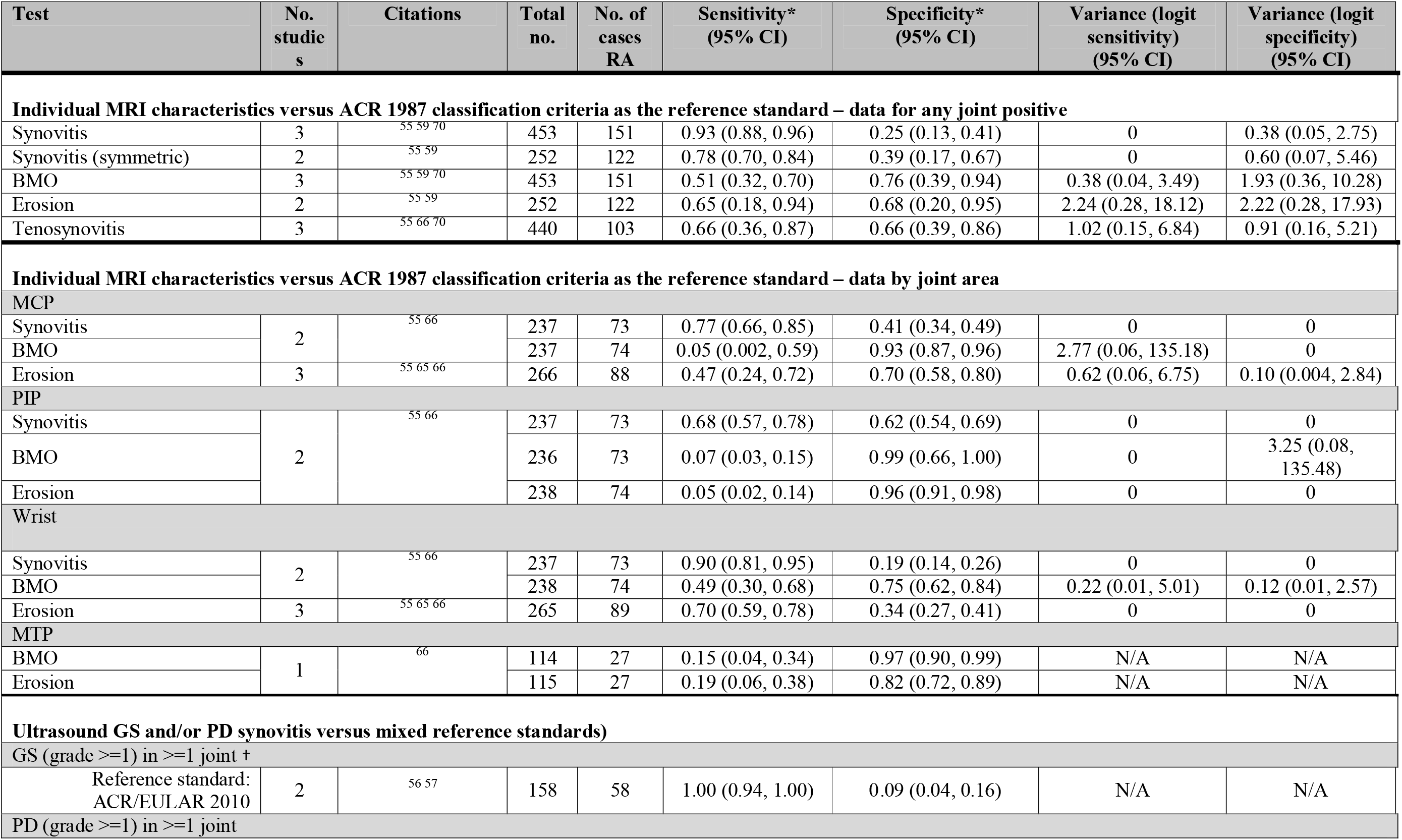
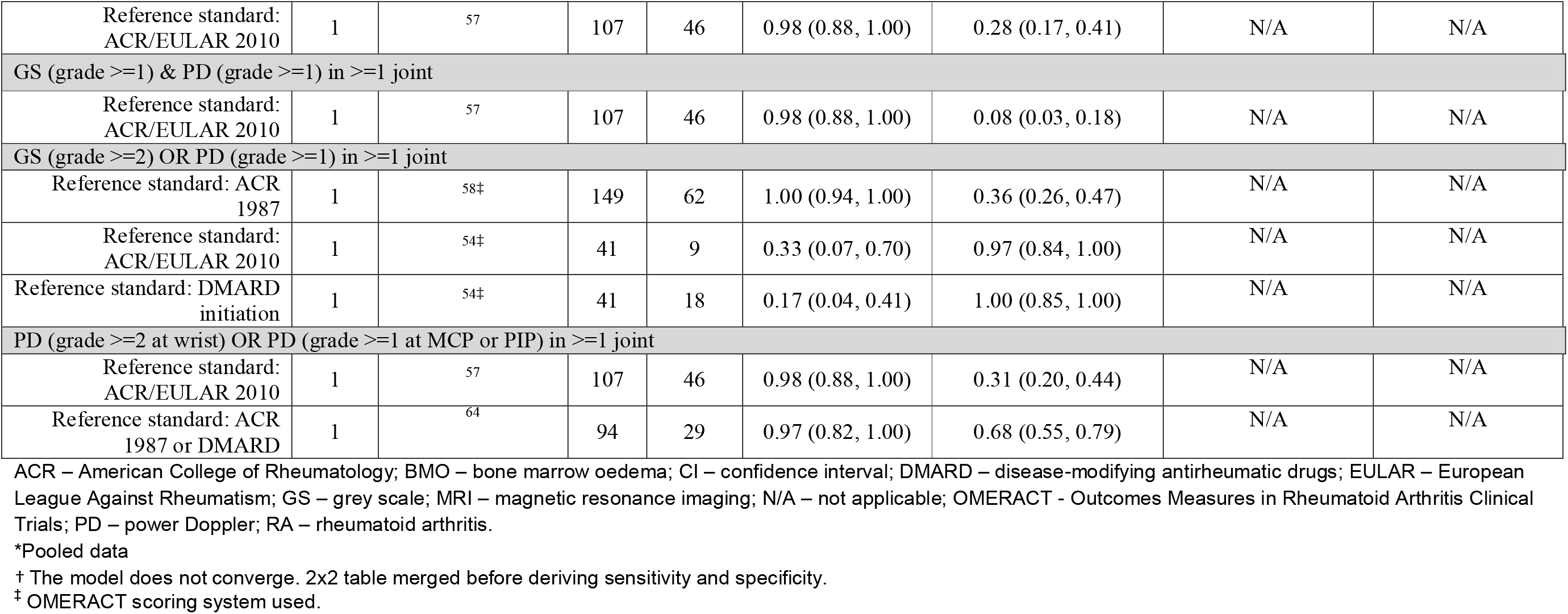
Summary of results for imaging test evaluations

Data for other MRI characteristics were heterogeneous but suggested lower sensitivities with some evidence of higher specificities (Figure 3). Summary sensitivities were 51% (95% CI 32% to 70%) for bone marrow oedema (BMO) (n=3^55 59 70^; 453 participants [151 with RA]), 65% (95% CI 18% to 94%) for erosion (n=2^55 59^; 252 [122]) and 66% (95% CI 36% to 87%) for tenosynovitis (n=3^55 66 70^; 440 [103 cases]). Respective summary specificities were 76% (95% CI 39% to 94%), 68% (95% CI 20% to 95%) and 66% (95% CI 39% to 86%) (Table 2).

A combination of MRI characteristics (synovitis/erosion, **or**, BMO and/or erosion) was assessed in three studies^59 60 67^, two using RAMRIS^59 67^ (Figure 4). Results were mixed, and data were insufficient to identify a clear effect on accuracy. Four studies^55 65–67^ presented results by joint area (MCP, PIP, wrist) using RAMRIS and ACR 1987 as reference (Table 2). Summary sensitivities were 47% to 77% and specificities 34% to 75%, however some characteristics were either highly sensitive or highly specific. Summary sensitivity for wrist synovitis was 90% (95% CI 81% to 95%)(n=2^55 66^) with specificity 19% (95% CI 14% to 26%). In contrast, BMO and erosion of PIPJ were highly specific (summary specificities 96% to 99%), with sensitivities 7% or less. Additional miscellaneous thresholds and reference standards data are presented in supplementary Figure 3.

**Figure 4.**
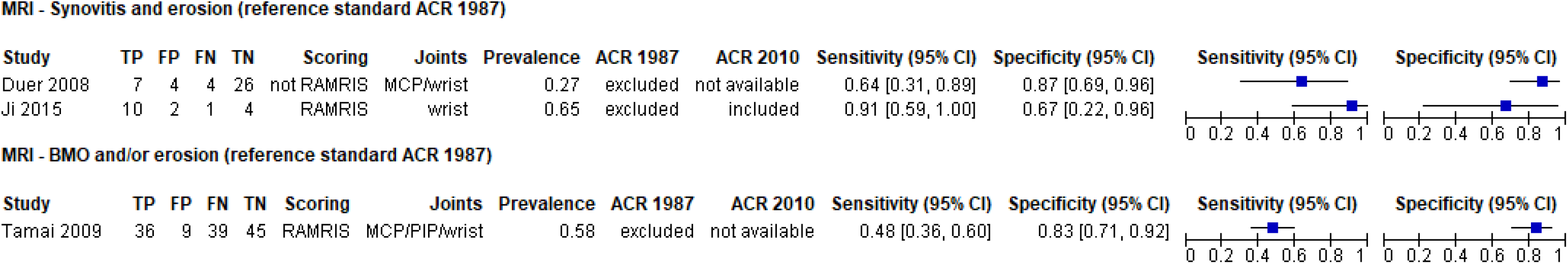
Forest plot of sensitivities and specificities from studies evaluating combinations of individual MRI characteristics (any joint positive against ACR 1987 reference standard)

#### Ultrasound performance

Ultrasound evaluations used a variety of scoring systems as reported above. Diagnostic thresholds and reference standards used varied, limiting statistical pooling.

Both GS and PD (grade >=1 in at least one joint) showed high sensitivities (98% or more) but low specificities (Table 2; Figure 5). One study^64^ reported higher specificities (up to 86%) for higher grades of synovitis, or for synovitis in greater number of joints (supplementary Figure 4). Three studies reporting combinations of GS and PD ^54 57 58^ had contrasting results with either high sensitivities ^57 58^ or high specificities^54^ (Table 2) with variations according to synovitis grade or number of joints (Figure 5; supplementary Figure 4). One study^57^ reported data by individual joints: specificities for joint synovitis (grade ≥1) were at least 90% for two PIP and one MTP joint on GS, and for three PIP (1 and 4-5) and three MTP (2-3 and 5) joints on PD (supplementary Figure 5); sensitivities were 37% or lower. The highest sensitivities were observed for wrist synovitis on both GS (89%, 95% CI 76, 96%) and PD (87%, 95% CI 74% to 95%)^57^.

**Figure 5.**
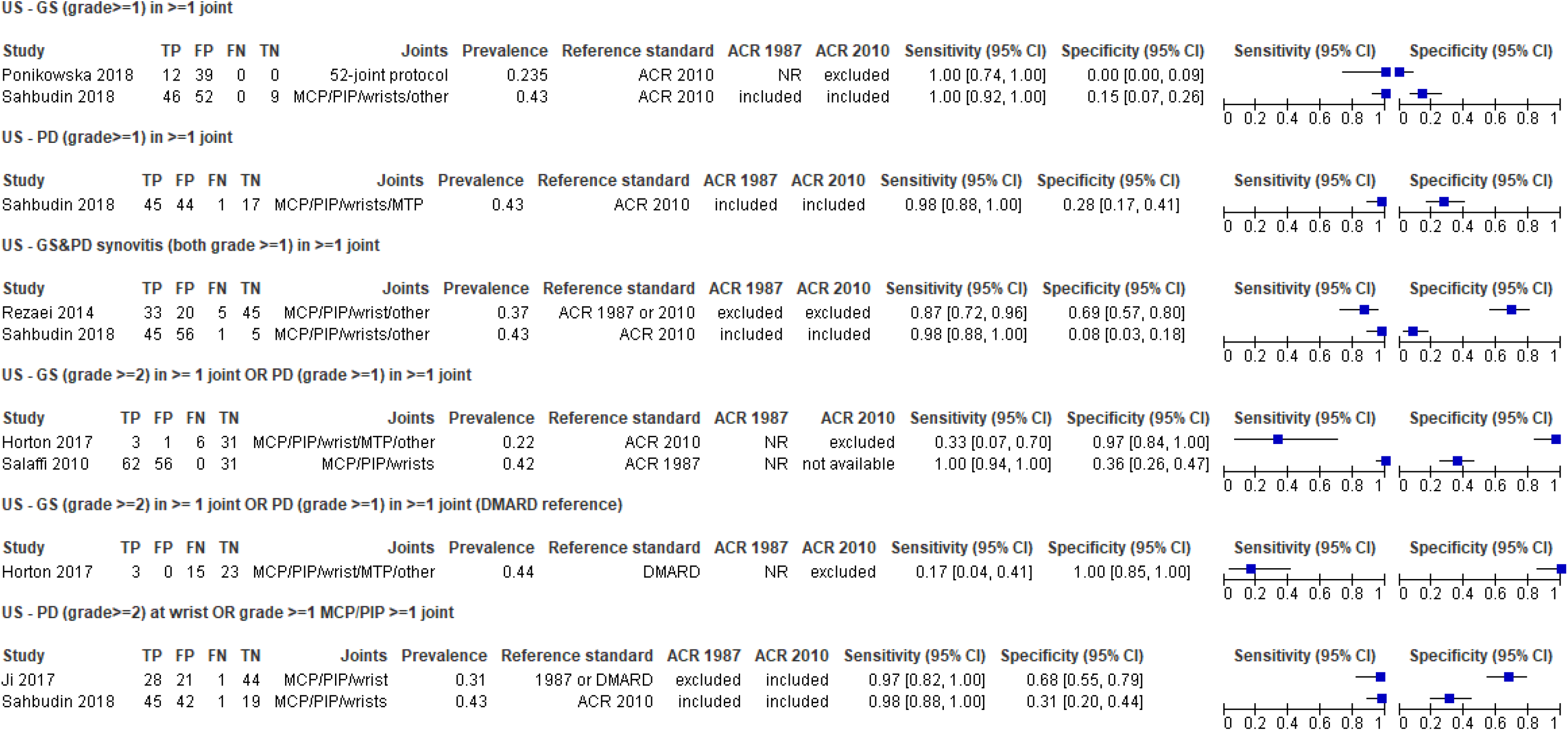
Forest plot of sensitivities and specificities from studies evaluating GS and/or PD ultrasound (any joint positive against mixed reference standards) * All ultrasound evaluations included both grey scale (GS) synovitis and power dower (PD) activity. Synovitis was scored using similar semiquantitative grading scales. ^24 48–50^ Two studies^53 57^ used OMERACT definition of synovitis^45 46^ and none used the EULAR-OMERACT combined scoring system for grading synovitis in RA^3–5^.

## DISCUSSION

We found 13 studies evaluating four MRI characteristics and 6 ultrasound studies evaluating synovitis. Studies were of variable methodological quality regarding both risk of bias and clinical applicability. Unclear participant recruitment procedures, potential inclusion of patients with RA at baseline, differential verification and lack of blinding were of particular concern. Only five studies included participants meeting our definition of UA^54 56 57 63 86^, others including at least some participants with tender joints but no synovitis^62–65,66–68^, or more than one joint with synovitis^60 61^ or including additional biomarkers^58,61,64 65^. Most MRI evaluations were blinded to the reference standard, but reported consensus or mean results across more than one observer so that results are unlikely to be replicable in a usual practice setting, in which one radiologist performs MRI interpretations. Most studies did however report using replicable scoring methods for both MRI and ultrasound.

Although the aim is earlier initiation of treatment to reduce morbidity and prevent irreversible damage in RA, of equal importance is to avoid inappropriate treatment with often expensive and potentially harmful treatments in those who do not need it. The ideal testing strategy in early disease should maximise sensitivity but not at the expense of specificity.

MRI synovitis had the highest and most homogenous sensitivities (summary sensitivity 93%, 95% CI 88% to 96%), however specificities, and therefore false positive rates, were poor and variable. The summary specificity of 25% (95% CI 13 to 41%) suggests as much as 75% of those who did not meet RA classification criteria also demonstrated synovitis. There was some evidence of higher specificities for other MRI characteristics, but studies were small and results inconsistent. Ultrasound results were difficult to synthesise due to different diagnostic thresholds, scoring systems and reference standards used.

For both tests, limited data suggested that combinations of findings from different joint areas could help to rule in/rule out those who are most/least likely to develop RA, however further work is needed to determine the optimal combination. Combining imaging test results with other clinical features and biomarkers in formal prediction rules may further add predictive power^58 59 63 66^ however this was not the focus of the current review.

### Challenges in evaluation

Due to varying definitions of UA and RA, we were unable to determine how MRI or ultrasound would perform in our target population of patients with UA who do not meet either set of RA classification criteria at the time of imaging. Almost all studies included people meeting at least one set of classification criteria at baseline; either explicitly reporting the exclusion of participants meeting only one set of criteria, or because the 2010 criteria had not yet been published. Tests may have performed differently if all of those with RA at baseline had been excluded. Changing classification criteria and small number of studies (n=4) using only the ACR/EULAR 2010 criteria as the reference standard was an additional challenge. Participants with resolving symptoms were poorly reported (5%-41%^63 57^, n=6).

There was a notable lack of consistency in ultrasound scoring. As expected, evaluations published prior to the EULAR-OMERACT combined scoring system for grading ultrasound synovitis in RA, used various ultrasound scoring systems^19 46 49 89–91 50 51 87 92 93^, but these have remained in use even in the most recent studies published after 2017. The potential for absence of subclinical inflammation to identify those most likely to resolve has not yet been considered.

Only three studies explicitly reported blinding of final disease status to the imaging test result and one reported lack of blinding to the imaging test. Although current classification criteria do not include imaging characteristics, knowledge of the imaging result may implicitly affect diagnosis.

### Strengths and weaknesses of this review

We used a comprehensive electronic literature search with stringent systematic review methods that included independent duplicate data extraction and quality assessment of studies, attempted contact with authors, and a clear analysis structure. We identified one previous systematic review^94^, that identified only two studies in UA populations (both of which were included in our review^59 60^) and included 11 studies in mixed populations, four of which we also included^61 65 68 69^. We excluded the other 7 studies on the basis of study population^95–98^, target condition^99^, lack of follow-up^100^, or lack of 2×2 data^101^. Limited statistical pooling was possible due to our stringent criteria for meta-analysis. Pooling was only considered for studies using similar imaging scoring systems and reference standards for RA. Although the 2010 criteria identify most patients meeting 1987 criteria (e.g. 91% of those meeting 1987 criteria in one study^102^), the 2010 criteria also classify considerably more patients as having RA compared to the 1987 criteria (e.g. in another study only 36% of those meeting 2010 criteria would have been diagnosed using the 1987 criteria^103^). The only study reporting accuracy against more than one reference standard^54^, reported RA prevalence 17% higher when defined by DMARD initiation compared to classification criteria definition.

### Implications for research

Future studies of imaging tests for the diagnosis of RA should be based on clearly defined UA populations (e.g. individuals with ≥1 swollen joint and who do not meet RA or other rheumatologic disease classification criteria at baseline). Symptoms and duration of symptoms should be specified. The reference standard diagnosis using current RA classification criteria should be made up to at least 12 months from baseline, and should be clearly blinded to imaging findings. Validated scoring methods and accepted thresholds for defining a positive test (e.g. RAMRIS^39 40^ and EULAR-OMERACT^42–44^ for MRI and US, respectively) should be used. Any future research study should conform to reporting guidelines, including the updated Standards for Reporting of Diagnostic Accuracy guideline^104^.

### Conclusion

There is currently insufficient evidence to recommend MRI or ultrasound alone for early RA diagnosis in people with UA. This is in large part because of a lack of consistency in study methodology, which also prevents synthesis of data in studies examining the combined utility of imaging and other clinical and serological tests. In order to address this, the research community needs to develop larger scale studies that apply consistent recruitment and scoring strategies and relate these to common reference standards, and that conform to international reporting guidelines for test accuracy studies ^36^.

## Data Availability

Available on request

## Acknowledgments

JD and PdeP contributed equally to this work. JD, PdeP, AO, SB and ZL undertook the review. JD, PdeP, KR, AF and JJD contributed to the conception of the work and interpretation of the findings. JD and PdeP drafted the manuscript. All authors critically revised the manuscript and approved the final version. PdeP acts as guarantor. The corresponding author attests that all listed authors meet authorship criteria and that no others meeting the criteria have been omitted.

We thank corresponding authors of included papers for providing additional data not contained in their publications, and Susan Bayliss for additional assistance with bibliographic search.

## Industry affiliations

None to report

The work has not been previously presented at a conference or meeting.

The lead authors and manuscript’s guarantor affirm that the manuscript is an honest, accurate and transparent account of the study being reported; that no important aspects of the study have been omitted; and that any discrepancies from the study as planned have been explained.

## References

1. Dadoun S, Zeboulon-Ktorza N, Combescure C, et al. Mortality in rheumatoid arthritis over the last fifty years: Systematic review and meta-analysis. Joint, bone, spine: revue du rhumatisme 2013;80(1):29–33. doi: https://doi.org/10.1016/j.jbspin.2012.02.005

2. Matcham F, Scott IC, Rayner L, et al. The impact of rheumatoid arthritis on quality-of-life assessed using the SF-36: A systematic review and meta-analysis. Seminars in Arthritis and Rheumatism 2014;44(2):123–30. doi: https://doi.org/10.1016/j.semarthrit.2014.05.001

3. Radner H, Smolen JS, Aletaha D. Comorbidity affects all domains of physical function and quality of life in patients with rheumatoid arthritis. Rheumatology 2010;50(2):381–88. doi: 10.1093/rheumatology/keq334

4. Lenssinck M-LB, Burdorf A, Boonen A, et al. Consequences of inflammatory arthritis for workplace productivity loss and sick leave: a systematic review. Annals of the Rheumatic Diseases 2013;72(4):493. doi: 10.1136/annrheumdis-2012-201998

5. Sullivan PW, Ghushchyan V, Huang X-Y, et al. Influence of Rheumatoid Arthritis on Employment, Function, and Productivity in a Nationally Representative Sample in the United States. The Journal of rheumatology 2010;37(3):544. doi: 10.3899/jrheum.081306

6. Tang K, Beaton DE, Gignac MAM, et al. The Work Instability Scale for rheumatoid arthritis predicts arthritis-related work transitions within 12 months. Arthritis Care & Research 2010;62(11):1578–87. doi: 10.1002/acr.20272

7. de Pablo P, Filer A, Raza K, et al. Does time matter in the management of rheumatoid arthritis? In: Royal College of Physicians. Horizons in Medicine. London: RCP, 2009.

8. Finckh A, Liang MH, van Herckenrode CM, et al. Long-term impact of early treatment on radiographic progression in rheumatoid arthritis: A meta-analysis. Arthritis Rheum 2006;55(6):864–72. doi: 10.1002/art.22353 [published Online First: 2006/12/02]

9. van der Linden MPM, le Cessie S, Raza K, et al. Long-term impact of delay in assessment of patients with early arthritis. Arthritis & Rheumatism 2010;62(12):3537–46. doi: https://doi.org/10.1002/art.27692

10. Bykerk VP, Hazes JM. When does rheumatoid arthritis start and can it be stopped before it does? Annals of the Rheumatic Diseases 2010;69(3):473. doi: 10.1136/ard.2009.116020

11. van Gaalen FA, Linn-Rasker SP, van Venrooij WJ, et al. Autoantibodies to cyclic citrullinated peptides predict progression to rheumatoid arthritis in patients with undifferentiated arthritis: a prospective cohort study. Arthritis Rheum 2004;50:709–15.

12. Aletaha D, Neogi T, Silman AJ, et al. 2010 rheumatoid arthritis classification criteria: an American College of Rheumatology/European League Against Rheumatism collaborative initiative. Ann Rheum Dis 2010;69(9):1580–8. doi: 10.1136/ard.2010.138461 [published Online First: 2010/08/12]

13. Arnett FC, Edworthy SM, Bloch DA, et al. The American Rheumatism Association 1987 revised criteria for the classification of rheumatoid arthritis. Arthritis Rheum 1988;31(3):315-24. [published Online First: 1988/03/01]

14. Cader MZ, Filer A, Hazlehurst J, et al. Performance of the 2010 ACR/EULAR criteria for rheumatoid arthritis: comparison with 1987 ACR criteria in a very early synovitis cohort. Annals of the Rheumatic Diseases 2011;70(6):949–55. doi: 10.1136/ard.2010.143560

15. El Miedany Y, Youssef S, Mehanna AN, et al. Development of a scoring system for assessment of outcome of early undifferentiated inflammatory synovitis. Joint, Bone, Spine: Revue du Rhumatisme 2008;75(2):155–62.

16. Van Der Helm-Van Mil AHM, Cessie SL, Van Dongen H, et al. A prediction rule for disease outcome in patients with recent-onset undifferentiated arthritis: How to guide individual treatment decisions. Arthritis and Rheumatism 2007;56(2):433–40.

17. Visser H, le Cessie S, Vos K, et al. How to diagnose rheumatoid arthritis early: A prediction model for persistent (erosive) arthritis. Arthritis & Rheumatism 2002;46(2):357–65. doi: 10.1002/art.10117

18. McQueen FM. Imaging in early rheumatoid arthritis. Best Practice & Research Clinical Rheumatology 2013;27(4):499–522. doi: https://doi.org/10.1016/j.berh.2013.09.005

19. Ohrndorf S, Backhaus M. Advances in sonographic scoring of rheumatoid arthritis. Ann Rheum Dis 2013;72 Suppl 2:ii69–75. doi: 10.1136/annrheumdis-2012-202197 [published Online First: 2012/12/21]

20. Emery P, van der Heijde D, Østergaard M, et al. Exploratory analyses of the association of MRI with clinical, laboratory and radiographic findings in patients with rheumatoid arthritis. Annals of the Rheumatic Diseases 2011;70(12):2126. doi: 10.1136/ard.2011.154500

21. Filer A, de Pablo P, Allen G, et al. Utility of ultrasound joint counts in the prediction of rheumatoid arthritis in patients with very early synovitis. Annals of the Rheumatic Diseases 2011;70(3):500–07.

22. American College of Rheumatology Rheumatoid Arthritis Clinical Trials Task Force Imaging Group, Outcome Measures in Rheumatology Magnetic Resonance Imaging Inflammatory Arthritis Working Group. Review: The Utility of Magnetic Resonance Imaging for Assessing Structural Damage in Randomized Controlled Trials in Rheumatoid Arthritis. Arthritis & Rheumatism 2013;65(10):2513–23. doi: 10.1002/art.38083

23. Hua C, Daien CI, Combe B, et al. Diagnosis, prognosis and classification of early arthritis: Results of a systematic review informing the 2016 update of the EULAR recommendations for the management of early arthritis. RMD Open 2017;3(1):e000406.

24. Cunnington J, Platt P, Raftery G, et al. Attitudes of United Kingdom rheumatologists to musculoskeletal ultrasound practice and training. Annals of the rheumatic diseases 2007;66(10):1381–83. doi: 10.1136/ard.2006.065466

25. Møller-Bisgaard S, Hørslev-Petersen K, Ejbjerg B, et al. Effect of Magnetic Resonance Imaging vs Conventional Treat-to-Target Strategies on Disease Activity Remission and Radiographic Progression in Rheumatoid Arthritis: The IMAGINE-RA Randomized Clinical Trial. JAMA 2019;321(5):461–72. doi: 10.1001/jama.2018.21362

26. Descamps L, Olagne L, Merlin C, et al. Utility of PET/CT in the diagnosis of inflammatory rheumatic diseases: A systematic review and meta-analysis. Annals of the Rheumatic Diseases 2018;77 (11) (no pagination)(e81)

27. Kubota K, Yamashita H, A. M. Clinical Value of FDG-PET/CT for the Evaluation of Rheumatic Diseases: Rheumatoid Arthritis, Polymyalgia Rheumatica, and Relapsing Polychondritis. Semin Nucl Med 2017;47:408–24.

28. Mattes-Gyorgy K, Buchbender C, Ostendorf B, et al. Synovitis and bone affection in early rheumatoid arthritis: High-resolution multi-pinhole single-photon-emission computed tomography (MPH-SPECT) versus magnetic resonance imaging (MRI). NuklearMedizin 2012;51 (2):A89–A90.

29. Thuermel K, Neumann J, Jungmann PM, et al. Fluorescence optical imaging and 3T-MRI for detection of synovitis in patients with rheumatoid arthritis in comparison to a composite standard of reference. European Journal of Radiology 2017;90:6–13.

30. Krohn M, Ohrndorf S, Werner SG, et al. Near-infrared Fluorescence Optical Imaging in Early Rheumatoid Arthritis: A Comparison to Magnetic Resonance Imaging and Ultrasonography. The Journal of rheumatology 2015;42:1112–18. doi: 10.3899/jrheum.141244

31. Werner SG, Langer HE, Schott P, et al. Indocyanine green-enhanced fluorescence optical imaging in patients with early and very early arthritis: a comparative study with magnetic resonance imaging. Arthritis Rheum 2013;65:3036–44.

32. Żuchowski P, Waszczak-Jeka M, Kudlicki S, et al. The rheumatoid hand in the light of fluorescence: a diagnostic technique of the future?. Reumatologia 2019;57:45–49. doi: 10.5114/reum.2019.83239

33. van Onna M, Ten Cate DF, Tsoi KL, et al. Assessment of disease activity in patients with rheumatoid arthritis using optical spectral transmission measurements, a non-invasive imaging technique. Annals of the Rheumatic Diseases 2016;75(3):511–18.

34. Meier AJL, Rensen W, de Bokx P, et al. Potential of optical spectral transmission measurements for joint inflammation measurements in rheumatoid arthritis patients. Journal of Biomedical Optics 2012;17(8):081420.

35. Deeks JJ, Bossuyt PM, C G, editors. Cochrane Handbook for Systematic Reviews of Diagnostic Test Accuracy Version 1.0.0.. Available from srdta.cochrane.org.: The Cochrane Collaboration, 2013.

36. McInnes MDF, Moher D, Thombs BD, et al. Preferred Reporting Items for a Systematic Review and Meta-analysis of Diagnostic Test Accuracy Studies: The PRISMA-DTA Statement. JAMA 2018;319(4):388–96. doi: 10.1001/jama.2017.19163

37. van Steenbergen HW, Aletaha D, Beaart-van de Voorde LJ, et al. EULAR definition of arthralgia suspicious for progression to rheumatoid arthritis. Ann Rheum Dis 2017;76:491–96.

38. Whiting PF, Rutjes AW, Westwood ME, et al. QUADAS-2: a revised tool for the quality assessment of diagnostic accuracy studies. Annals of internal medicine 2011;155(8):529–36. doi: 10.7326/0003-4819-155-8-201110180-00009 [published Online First: 2011/10/19]

39. Haavardsholm EA, Ostergaard M, Ejbjerg BJ, et al. Introduction of a novel magnetic resonance imaging tenosynovitis score for rheumatoid arthritis: reliability in a multireader longitudinal study. Ann Rheum Dis 2007;66(9):1216–20. doi: 10.1136/ard.2006.068361 [published Online First: 2007/03/30]

40. Ostergaard M, Peterfy C, Conaghan P, et al. OMERACT Rheumatoid Arthritis Magnetic Resonance Imaging Studies. Core set of MRI acquisitions, joint pathology definitions, and the OMERACT RA-MRI scoring system. The Journal of rheumatology 2003;30(6):1385-6. [published Online First: 2003/06/05]

41. Szkudlarek M, Court-Payen M, Jacobsen S, et al. Interobserver agreement in ultrasonography of the finger and toe joints in rheumatoid arthritis. Arthritis Rheum 2003;48(4):955–62. doi: 10.1002/art.10877 [published Online First: 2003/04/11]

42. Bruyn GA, Iagnocco A, Naredo E, et al. OMERACT Definitions for Ultrasonographic Pathologies and Elementary Lesions of Rheumatic Disorders 15 Years On. The Journal of rheumatology 2019;46(10):1388–93. doi: 10.3899/jrheum.181095

43. D’Agostino M-A, Terslev L, Aegerter P, et al. Scoring ultrasound synovitis in rheumatoid arthritis: a EULAR-OMERACT ultrasound taskforce<strong>—</strong>Part 1: definition and development of a standardised, consensus-based scoring system. RMD Open 2017;3(1):e000428. doi: 10.1136/rmdopen-2016-000428

44. Terslev L, Naredo E, Aegerter P, et al. Scoring ultrasound synovitis in rheumatoid arthritis: a EULAR-OMERACT ultrasound taskforce-Part 2: reliability and application to multiple joints of a standardised consensus-based scoring system. RMD Open 2017;3(1):e000427. doi: 10.1136/rmdopen-2016-000427

45. Wakefield RJ, Balint PV, Szkudlarek M, et al. Musculoskeletal ultrasound including definitions for ultrasonographic pathology. The Journal of rheumatology 2005;32(12):2485-7. [published Online First: 2005/12/07]

46. Naredo E, Wakefield RJ, Iagnocco A, et al. The OMERACT Ultrasound Task Force — Status and Perspectives. The Journal of rheumatology 2011;38(9):2063. doi: 10.3899/jrheum.110425

47. Naredo E, D’Agostino MA, Wakefield RJ, et al. Reliability of a consensus-based ultrasound score for tenosynovitis in rheumatoid arthritis. Ann Rheum Dis 2013;72(8):1328–34. doi: 10.1136/annrheumdis-2012-202092 [published Online First: 2012/09/18]

48. Boers M, Brooks P, Strand CV, et al. The OMERACT filter for Outcome Measures in Rheumatology. The Journal of rheumatology 1998;25(2):198-9. [published Online First: 1998/03/07]

49. Szkudlarek M, Court-Payen M, Jacobsen S, et al. Interobserver agreement in ultrasonography of the finger and toe joints in rheumatoid arthritis. Arthritis Rheum 2003;48(4):955–62. doi: 10.1002/art.10877 [published Online First: 2003/04/11]

50. Schmidt WA, Schmidt H, Schicke B, et al. Standard reference values for musculoskeletal ultrasonography. Ann Rheum Dis 2004;63(8):988–94. doi: 10.1136/ard.2003.015081 [published Online First: 2004/07/14]

51. Szkudlarek M, Klarlund M, Narvestad E, et al. Ultrasonography of the metacarpophalangeal and proximal interphalangeal joints in rheumatoid arthritis: a comparison with magnetic resonance imaging, conventional radiography and clinical examination. Arthritis Res Ther 2006;8(2):R52. doi: 10.1186/ar1904 [published Online First: 2006/03/08]

52. Reitsma JB, Glas AS, Rutjes AWS, et al. Bivariate analysis of sensitivity and specificity produces informative summary measures in diagnostic reviews. Journal of Clinical Epidemiology 2005;58(10):982–90. doi: https://doi.org/10.1016/j.jclinepi.2005.02.022

53. Takwoingi Y, Guo B, Riley RD, et al. Performance of methods for meta-analysis of diagnostic test accuracy with few studies or sparse data. Stat Methods Med Res 2017;26(4):1896–911. doi: 10.1177/0962280215592269 [published Online First: 2015/06/28]

54. Horton SC, Tan AL, Wakefield RJ, et al. Ultrasound-detectable grey scale synovitis predicts future fulfilment of the 2010 ACR/EULAR RA classification criteria in patients with new-onset undifferentiated arthritis. RMD Open 2017;3(1):e000394.

55. Li R, Liu X, Ye H, et al. Magnetic resonance imaging in early rheumatoid arthritis: a multicenter, prospective study. Clinical Rheumatology 2016;35(2):303–8.

56. Ponikowska M, Swierkot J, Nowak B. The importance of ultrasound examination in early arthritis. Reumatologia 2018;56(6):354–61. doi: http://dx.doi.org/10.5114/reum.2018.80712

57. Sahbudin I, Pickup L, Nightingale P, et al. The role of ultrasound-defined tenosynovitis and synovitis in the prediction of rheumatoid arthritis development. Rheumatology (Oxford) 2018;57(7):1243–52.

58. Salaffi F, Ciapetti A, Gasparini S, et al. A clinical prediction rule combining routine assessment and power Doppler ultrasonography for predicting progression to rheumatoid arthritis from early-onset undifferentiated arthritis. Clinical & Experimental Rheumatology 2010;28(5):686– 94.

59. Tamai M, Kawakami A, Uetani M, et al. A prediction rule for disease outcome in patients with undifferentiated arthritis using magnetic resonance imaging of the wrists and finger joints and serologic autoantibodies. Arthritis & Rheumatism 2009;61(6):772–8.

60. Duer A, Ostergaard M, Horslev-Petersen K, et al. Magnetic resonance imaging and bone scintigraphy in the differential diagnosis of unclassified arthritis. Annals of the Rheumatic Diseases 2008;67(1):48–51.

61. Narvaez J, Sirvent E, Narvaez JA, et al. Usefulness of magnetic resonance imaging of the hand versus anticyclic citrullinated peptide antibody testing to confirm the diagnosis of clinically suspected early rheumatoid arthritis in the absence of rheumatoid factor and radiographic erosions. Seminars in Arthritis & Rheumatism 2008;38(2):101–9.

62. Aoki T, Yamashita Y, Saito K, et al. Diagnosis of early-stage rheumatoid arthritis: usefulness of unenhanced and gadolinium-enhanced MR images at 3 T. Clinical Imaging 2013;37(2):348– 53.

63. Lei X, Li H, Zhan Y, et al. Predict rheumatoid arthritis conversion from undifferentiated arthritis with dynamic contrast-enhanced MRI and laboratory indexes. Clinical & Experimental Rheumatology 2018;36(4):552–58.

64. Ji L, Deng X, Geng Y, et al. The additional benefit of ultrasonography to 2010 ACR/EULAR classification criteria when diagnosing rheumatoid arthritis in the absence of anti-cyclic citrullinated peptide antibodies. Clinical Rheumatology 2017;36(2):261–67.

65. Solau-Gervais E, Legrand JL, Cortet B, et al. Magnetic resonance imaging of the hand for the diagnosis of rheumatoid arthritis in the absence of anti-cyclic citrullinated peptide antibodies: a prospective study. Journal of Rheumatology 2006;33(9):1760–5.

66. Duer-Jensen A, Horslev-Petersen K, Hetland ML, et al. Bone edema on magnetic resonance imaging is an independent predictor of rheumatoid arthritis development in patients with early undifferentiated arthritis. Arthritis & Rheumatism 2011;63(8):2192–202.

67. Ji L, Li G, Xu Y, et al. Early prediction of rheumatoid arthritis by magnetic resonance imaging in the absence of anti-cyclic citrullinated peptide antibodies and radiographic erosions in undifferentiated inflammatory arthritis patients: a prospective study. International Journal of Rheumatic Diseases 2015;18(8):859–65.

68. Klarlund M, Ostergaard M, Jensen KE, et al. Magnetic resonance imaging, radiography, and scintigraphy of the finger joints: One year follow up of patients with early arthritis. Annals of the Rheumatic Diseases 2000;59(7):521–28.

69. Mori G, Tokunaga D, Takahashi KA, et al. Maximum intensity projection as a tool to diagnose early rheumatoid arthritis. Modern Rheumatology 2008;18(3):247–51.

70. Boer AC, Burgers LE, Mangnus L, et al. Using a reference when defining an abnormal MRI reduces false-positive MRI results-a longitudinal study in two cohorts at risk for rheumatoid arthritis. Rheumatology (Oxford) 2017;56(10):1700–06.

71. Boeters DM, Boer AC, Van Der Helm-Van Mil AHM. Evaluation of the predictive accuracy of MRI-detected erosions in hand and foot joints in patients with undifferentiated arthritis. Annals of the Rheumatic Diseases 2019;78(1):144–46.

72. Rezaei H, Torp-Pedersen S, af Klint E, et al. Diagnostic utility of musculoskeletal ultrasound in patients with suspected arthritis--a probabilistic approach. Arthritis Research & Therapy 2014;16(5):448.

73. Boer AC, Boeters DM, Van Der Helm-Van Mil AHM. The use of MRI-detected synovitis to determine the number of involved joints for the 2010 ACR/EULAR classification criteria for Rheumatoid Arthritis-is it of additional benefit? Annals of the Rheumatic Diseases 2018;77(8):1125–29.

74. Nieuwenhuis WP, Reijnierse M, van der Helm-van Mil AH. Does adding the presence of MRI detected bone marrow oedema improve the accuracy of the 2010 EULAR/ACR criteria for rheumatoid arthritis? Annals of the Rheumatic Diseases 2015;74(3):e29.

75. Chaiamnuay S, Lopez-Ben R, Alarcon GS. Ultrasound of target joints for the evaluation of possible inflammatory arthropathy: associated clinical factors and diagnostic accuracy. Clinical & Experimental Rheumatology 2008;26(5):875–80.

76. Mayordomo L, Jurado C, Almeida C, et al. PREDICTIVE VALUE OF ULTRASONOGRAPHY FOR RADIOGRAPHIC PROGRESION IN PATIENTS WITH EARLY RHEUMATOID ARTHRITIS. Annals of the Rheumatic Diseases 2016;75(Suppl. 2):868–69.

77. Mayordomo L, Jurado C, Velloso ML, et al. SERONEGATIVE EARLY ARTHRITIS. MAY POWER DOPPLER ULTRASONOGRAPHY PREDICT PROGRESSION TO RHEUMATOID ARTHRITIS? Annals of the Rheumatic Diseases 2016;75(Suppl. 2):1228–29.

78. Mayordomo L, Velloso ML, Almeida C, et al. PREDICTIVE VALUE OF POWER DOPPLER ULTRASONOGRAPHY (PDUS) FOR THE DIAGNOSIS OF RHEUMATOID ARTHRITIS IN PATIENTS WITH BASAL NEGATIVE ACUTE PHASE REACTANTS. Annals of the Rheumatic Diseases 2016;75(Suppl. 2):868.

79. Mayordomo L, Jurado C, Velloso ML, et al. Predictive value of power doppler ultrasonography (PDUS) in the diagnosis of early rheumatoid arthritis. Annals of the Rheumatic Diseases 2017;76 (Supplement 2):728.

80. Mayordomo L, Velloso ML, Jurado C, et al. Value of power doppler ultrasonography for prediction of treatment for rheumatoid arthritis (RA) in early arthritis during the first 12 monts of follow-up. Annals of the Rheumatic Diseases 2017;76 (Supplement 2):1018.

81. Mayordomo L, Almeida C, Jurado MC, et al. ULTRASONOGRAPHY POWER DOPPLER(PDUS) IN EARLY ARTHRITIS. DOES 44 JOINT COUNT PREDICTE MORE ACCURETLY THE DEVELOPMENT OF RA THAN OTHER ULTRASOUND COUNTS? Annals of the Rheumatic Diseases 2018;77(Suppl. 2):1700.

82. Nieuwenhuis WP, Krabben A, Stomp W, et al. Evaluation of magnetic resonance imaging-detected tenosynovitis in the hand and wrist in early arthritis. Arthritis & Rheumatology 2015;67(4):869–76.

83. Nieuwenhuis WP, van Steenbergen HW, Mangnus L, et al. Evaluation of the diagnostic accuracy of hand and foot MRI for early Rheumatoid Arthritis. Rheumatology 2017;56(8):1367–77. doi: 10.1093/rheumatology/kex167

84. Tavares R, Beattie KA, Bensen WG, et al. A double-blind, randomized controlled trial to compare the effect of biannual peripheral magnetic resonance imaging, radiography and standard of care disease progression monitoring on pharmacotherapeutic escalation in rheumatoid and undifferentiated inflammatory arthritis: study protocol for a randomized controlled trial. Trials [Electronic Resource] 2014;15:268.

85. Boer AC, Burgers LE, Mangnus L, et al. Evaluation of the accuracy of hand and foot MRI in the early identification of RA: Using the prevalence of low-graded inflammation in the symptom-free population as reference reduces false-positive MRI results. Annals of the Rheumatic Diseases 2017;76 (Supplement 2):473–74.

86. Li H, Qu J, Zhan Y, et al. Predictive value of dynamic enhanced MR synovial wash in rate in progression to rheumatoid arthritis in patients with undifferentiated arthritis. [Chinese]. National Medical Journal of China 2016;96(41):3315–18.

87. Backhaus M, Ohrndorf S, Kellner H, et al. Evaluation of a novel 7-joint ultrasound score in daily rheumatologic practice: a pilot project. Arthritis Rheum 2009;61(9):1194–201. doi: 10.1002/art.24646 [published Online First: 2009/08/29]

88. Klarlund M, Ostergaard M, Rostrup E, et al. Dynamic magnetic resonance imaging of the metacarpophalangeal joints in rheumatoid arthritis, early unclassified polyarthritis, and healthy controls. Scandinavian Journal of Rheumatology 2000;29(2):108–15.

89. Nakagomi D, Ikeda K, Okubo A, et al. Ultrasound can improve the accuracy of the 2010 American College of Rheumatology/European League against rheumatism classification criteria for rheumatoid arthritis to predict the requirement for methotrexate treatment. Arthritis Rheum 2013;65(4):890–8. doi: 10.1002/art.37848 [published Online First: 2013/01/22]

90. Ceponis A, Onishi M, Bluestein HG, et al. Utility of the ultrasound examination of the hand and wrist joints in the management of established rheumatoid arthritis. Arthritis Care Res (Hoboken) 2014;66(2):236–44. doi: 10.1002/acr.22119 [published Online First: 2013/08/29]

91. Nam JL, Hensor EM, Hunt L, et al. Ultrasound findings predict progression to inflammatory arthritis in anti-CCP antibody-positive patients without clinical synovitis. Ann Rheum Dis 2016;75(12):2060–67. doi: 10.1136/annrheumdis-2015-208235 [published Online First: 2016/01/24]

92. Szkudlarek M, Court-Payen M, Strandberg C, et al. Power Doppler ultrasonography for assessment of synovitis in the metacarpophalangeal joints of patients with rheumatoid arthritis: a comparison with dynamic magnetic resonance imaging. Arthritis Rheum 2001;44(9):2018–23. doi: 10.1002/1529-0131(200109)44:9<2018::Aid-art350>3.0.Co;2-c [published Online First: 2001/10/11]

93. Kitchen J, Kane D. Greyscale and power Doppler ultrasonographic evaluation of normal synovial joints: correlation with pro- and anti-inflammatory cytokines and angiogenic factors. Rheumatology (Oxford) 2015;54(3):458–62. doi: 10.1093/rheumatology/keu354 [published Online First: 2014/09/07]

94. Machado PM, Koevoets R, Bombardier C, et al. The value of magnetic resonance imaging and ultrasound in undifferentiated arthritis: a systematic review. Journal of Rheumatology - Supplement 2011;87:31–7.

95. Sugimoto H, Takeda A, Masuyama JI, et al. Early-stage rheumatoid arthritis: Diagnostic accuracy of MR imaging. Radiology 1996;198(1):185–92.

96. Sugimoto H, Takeda A, Hyodoh K. Early-stage rheumatoid arthritis: prospective study of the effectiveness of MR imaging for diagnosis. Radiology 2000;216(2):569–75.

97. Scire CA, Montecucco C, Codullo V, et al. Ultrasonographic evaluation of joint involvement in early rheumatoid arthritis in clinical remission: power Doppler signal predicts short-term relapse. Rheumatology 2009;48(9):1092–7.

98. Tamai M, Kawakami A, Uetani M, et al. Early prediction of rheumatoid arthritis by serological variables and magnetic resonance imaging of the wrists and finger joints: results from prospective clinical examination. Annals of the Rheumatic Diseases 2006;65(1):134–5.

99. Freeston JE, Wakefield RJ, Conaghan PG, et al. A diagnostic algorithm for persistence of very early inflammatory arthritis: the utility of power Doppler ultrasound when added to conventional assessment tools.[Erratum appears in Ann Rheum Dis. 2011 Aug;70(8):1519]. Annals of the Rheumatic Diseases 2010;69(2):417–9.

100. Boutry N, Hachulla E, Flipo RM, et al. MR imaging findings in hands in early rheumatoid arthritis: comparison with those in systemic lupus erythematosus and primary Sjogren syndrome. Radiology 2005;236(2):593–600.

101. Zampogna G, Parodi M, Bartolini B, et al. Dynamic contrast-enhanced magnetic resonance imaging of the wrist in early arthritis. [Italian]. Reumatismo 2008;60(4):254–59.

102. Jung SJ, Lee S-W, Ha YJ, et al. Patients with early arthritis who fulfil the 1987 ACR classification criteria for rheumatoid arthritis but not the 2010 ACR/EULAR criteria. Annals of the Rheumatic Diseases 2012;71(6):1097–98. doi: 10.1136/annrheumdis-2011-200785

103. Aga A-B, Lie E, Uhlig T, et al. In an Inception Cohort of 175 DMARD NaÏve RA Patients Classified According to the 2010 ACR/EULAR Criteria a Large Proportion of Patients does not Fulfill the 1987 ACR Criteria. Annals of the Rheumatic Diseases 2013;72(Suppl 3):A334–A34. doi: 10.1136/annrheumdis-2013-eular.1034

104. Bossuyt PM, Reitsma JB, Bruns DE, et al. STARD 2015: An Updated List of Essential Items for Reporting Diagnostic Accuracy Studies. Clinical Chemistry 2015;61(12):1446–52. doi: 10.1373/clinchem.2015.246280

