## Supplementary data for "Systematic review of imaging tests to predict the development of rheumatoid arthritis in people with unclassified arthritis"

**Supplementary information**

### Supplementary Table 1 - Summary characteristics of studies

| **Study**  **Sample size**  **(no. excluded)** | **UA definition;**  **maximum symptom duration (median; range)** | **RA at baseline** | | | **Sample characteristics** | | | | **Index test and interpretation** | | **Reference standard; FU**  **Prevalence (RA and other diagnoses)** |
| --- | --- | --- | --- | --- | --- | --- | --- | --- | --- | --- | --- |
|  |  | **ACR 1987** | **ACR 2010** | | **Female (n, %)**  **Mean age** | **ACPA +ve**  **RF +ve**  **(n, %)** | **SJC**  **TJC**  **median (range)** | **DMARD**  **during study** | **Tesla; joints imaged** | **Observers** |  |
| **MRI – STUDIES EVALUATING INDIVIDUAL CHARACTERISTICS (n=13 studies)** | | | | | | | | | | | |
| **Aoki 2013 ^1^**  N = 41 (4 exclusions) | Polyarthralgia incl wrist & <4 ACR 1987 criteria; NR (NR) | Excl | NR | | 36, 88%  59y | NR  16, 43% | NR  NR | NR | 3.0T; wrist (most symptomatic), hands (both) | 2 rheumatologists (>10y exp) | ACR 1987; mean 29.3 mos (12.2 to 45.5 mos)  RA 21, 57%; other confirmed dx 12, 32%; UA 4, 11% |
| **Boer 2017 ^2^**  N = 201 | NR ('clinically confirmed' arthritis);  < 2y (NR) | NR | Excl* | | 123, 61%  54y | 8, 4%  19, 10% | 2 (IQR 1, 4)  3 (IQR 1, 6) | 75, 37% | 1.5T; MCP, wrist, MTP (most affected or dominant) | NR; 2 of 3 ‘trained’ observers | ACR 1987; min 12 mos  RA 29, 14%; not RA 172, 86% |
| **Boeters 2019 ^3^**  N = 286 | NR ('clinically confirmed' UA);  < 2y (8 wks; 5-25 wks) | NR | Excl | | 174, 61%  55y | 24, 8%  32, 11% | 2 (IQR 1, 4)  3 (IQR 1, 7) | 111, 39%‡ | 1.5T; MCP, wrist, MTP (most affected or dominant) | NR; ‘2 observers’ | ACR 2010 (n=17) or clin dx + DMARD (n=111); min 12 mos  RA 128, 45%; UA 158, 55% |
| **Duer 2008 ^4^**  N = 41 | synovitis >=2 joints & pain or swelling of hand;  >6 mos (18 mos; 6 mos – 15y) | Excl | N/A | | 35, 85%  55y | NR  NR | 4 (2, 18)  6 (0, 30) | NR | 0.2T; MCP, wrist (most symptomatic) | NR | ACR 1987; min 24 mos  RA 11, 27%; not RA 30, 73% |
| **Duer-Jensen 2011 ^5^**  N = 127 (11 excluded) | synovitis >= 2 joints &/or >=2 tender joints;  6 mos to < 2y (RA 4 mos; 2-18 mos. Non-RA 7 mos; 18-24 mos) | Excl | N/A | | 90, 78%  RA; non-RA  50y; 47y | NR  NR | 11 (0, 34)  7 (0, 36) | NR | 0.6T or 1.0T; MTP, wrist, MCP, PIP (dominant) | 1 of 2 experienced rheumatologists | ACR 1987; median 17 mos, (12–23 mos)  RA 27, 24%; other confirmed dx 12, 11%; UA 28, 25%; other 49, 43% (athralgia/joint tenderness 34; self-limiting disease 15) |
| **Ji 2015 ^6^**  N = 31 | ≥2 tender &/or ≥2 swollen joints (MCP or wrist);  6 wks to ≤ 2y (12.3 mos SD 7.8 mos (RA), 8.3 mos (non-RA)) | Excl | Incl (14, 45%) | | 23, 74.2%  RA; non-RA  51y; 49y | NR  5, 16% | NR  NR | NR | 3.0T; both hands | 2 experienced radiologists | ACR 1987; median 15 mos (12-20 mos)  RA 22, 71%; other confirmed dx 6, 19%; UA 3, 10% |
| **Klarlund 2000 ^7^**  N = 24 (11 excluded) † | symmetric synovitis or tender MCP2-3 or PIP2-3;  < 2y (RA 3 mos; 2-5 mos. Non-RA 3.5 mos 1-13 mos) | Excl | N/A | | 12, 92%  40y§ | NR  4/13, 31% | RA; UA  3 (0, 8); 4 (0, 11)  13 (8, 20); 15 (2, 24) | 5/13, 38% | 1.0T; MCP dominant hand | single rheumatologist | ACR 1987; min 12 mos  RA 5, 36%; UA 8, 64% |
| **Lei 2018 ^8^**  N = 81 | synovitis >=1 joint or pain >= 1 joint of wrist and hands;  <= 1y (3.2 mos; 7 days – 3 mos) | NR | Excl | | 65, 80%  51y | 18, 22%  15, 19% | NR; 'joints involved'  RA; UA  6 (2, 18); 4 (2, 20) | 37, 46%‡ | 3.0T; wrist, hands (both) | single board certified radiologist | ACR 2010; min 12 mos  RA 37, 46%; other confirmed dx 28, 35%; UA 12, 15%; resolving 4, 5% |
| **Li 2016 ^9^**  N = 129 (6 excluded) | synovitis >=1 joint & athralgia >=1 of MCP/PIP/wrist;  <= 1y (mean 5 mos; 0.33-12 mos) | NR | Incl | | 93, 72%  47y | NR  NR | NR; symmetric arthritis: 113, 92%; arthritis >=3 joint areas: 76, 62% | NR | 1.5T; wrist, MCP, PIP (both) | NR; ‘single reader’ | ACR 1987; min 12 mos  RA 47, 38%; other confirmed dx 72, 59%; UA 4, 3% |
| **Mori 2008 ^10^**  N = 21 (4 excluded) | ‘undiagnosed arthritis of the hands’;  NR (NR) | Excl | N/A | | 14, 82%  58y | NR  NR | RA; non-A  1 SD 0.7 (0, 2); 3.2 SD 3.2 (0, 12)  NR | NR | 1.5T; 22 joints hands and distal forearms (both) | 2 rheumatologists | ACR 1987; mean 27.4 mos (13-40 mos)  RA 5 29%; not RA 7, 71% |
| **Narvaez 2008 ^11^**  N = 40 | RF –ve, symmetric arthritis >=3 joints (including >=1 wrist, MCP or PIP) & morning stiffness >=30 mins;  >=6 wks (4 mos; SD 2.6 mos) | Excl | N/A | | 28, 70%  54y | 7, 18%  0, 0% | mean 8 (SD 4)  mean 12 (SD 5) | 31, 77%‡ | 1.5T; wrist, MCP2-5 and PIP 2-5 (most affected) | 1 experienced radiologist | ACR 1987+DMARD (n=21) or DMARD alone (n=10); mean 20 mos (12-42 mos)  RA: 31, 77%; other confirmed dx 2, 5%; UA 7, 18% |
| **Solau-Gervais 2006 ^12^**  N = 30 (2 to 4 excluded) | symmetric polyarthritis or polyarthralgia of wrists and MCP &  morning stiffness ≥ 45 minutes +/- synovitis & ACPA negative;  < 2 y (mean 7.8 mos; SD 6.21 y) | Incl | N/A | | NR  46.8y | 0, 0%  10, 33% | mean 2.03 (0, 7)  mean 7.1 (SD 5.7) | 15, 50%‡ | 1.5T; wrist, MCP (both) | 2 rheumatologists | ACR 1987; mean 30.6 mos, min 12 mos  RA 16, 53%; other confirmed dx 10, 33%; UA 4, 13% |
| **Tamai 2009 ^13^**  N = 129 | synovitis wrists/fingers;  NR (3 mos; RA 0.5-15 mos; no RA 0.5-24 mos) | Excl | NR | | 100, 78%  RA; non-RA 53 y§; 52 y§ | NR  NR | RA; non-RA  3 (0, 23); 0 (0, 22)  6 (0, 28); 4 (0, 24) | 66, 51% | 1.5T; wrist, CMC1, CMC 2-5 joints, MCP1-5, and PIP1-5 (both) | 2 experienced radiologists | ACR 1987; min 12 mos  RA 75, 58%; other confirmed dx 35, 27%; UA 13, 10%; other 6, 5% (chronic hepatitis) |
| **US – STUDIES EVALUATING GS +/- PD (n=6 studies)** | | | | | | | | | | | |
| **Horton 2017 ^14^**  N = 60 (19 excluded) | synovitis >=1 joint; NR ‘new onset’ (9 mos; IQR 4-18 mos) | NR | | Excl | 27/41, 66%  45 y | 3, 7%  NR | 2 (IQR 1, 5)  NR | 45, 75%‡  (18/41 final sample) | GS&PD; 26 joint protocol (elbows, wrists, MCP2-3, PIP, knees, ankles, MTP1-5) | NR; presume consultant rheumatologist | ACR 2010; DMARD; min 12 mos  RA 9, 22% (ACR); other 32, 78% (dx NR for subgroup with US data - Of the 60 originally recruited: 13 (22%) developed RA, 32 (53%) UA, 2 (3%) other confirmed dx, 15 (25%) resolving) |
| **Ji 2017 ^15^**  N = 94 | ≥1 tender &/or swollen joint (hand) & pain &/or morning stiffness >30 minutes) & anti-CCP-ve; NR (RA 12.6 mos; SD 11.6 mos. non-RA 10.0 mos; SD 9.7 mos) | Excl | | Incl (30, 32%) | 48, 51%  RA; non-RA  58 y§; 51 y§ | 0, 0%**  11, 12% | RA; non-RA  4 (IQR 8); 1 (IQR 4); >=2 swollen joints: 51, 45%  10 (IQR 11); 5 (IQR 8); >=2 tender joints: 61, 36% | ?30, 31% | GS&PD; 22 joint protocol (wrists, MCP1-5, PIP1-5) | single trained rheumatologist | ACR 1987^\|^ ;median 5 mos (12-20 mos);  RA 29, 31%; other confirmed dx 47, 50%; UA 12, 12.8%; other (NR) 6, 6% |
| **Ponikowska 2018 ^16^**  N = 51† | oedema >=1 joint; < 1y (mean 5.9 mos; SD 3.9) | NR | | Excl | 37, 72%  47y | 2, 4%  8, 16% | mean 6 (SD 4) (0, 17)  mean 5.9 (SD 4.4) (0, 18) | NR‡ | GS¶; 52 joint protocol; MCP1-5, PIP2-5, DIP1-5, tarsal joints, MTP1-5, IP joint, PIP(toes) | physician with 10-year experience in MSK US | ACR 2010; min 12 mos  RA 12, 32%; other confirmed dx 16, 32%; UA 15, 30%; resolving 7, 14% |
| **Rezaei 2014 ^17^**  N = 103 | Inflammatory symptoms including ‘arthralgia, stiffness and swollen joints, mostly of the hands and feet'; NR (mean 8.5 mos; SD 3.8 mos) | Excl | | Excl | 76, 74%  50 y | 30, 29%  36, 34% | NR  NR | 55, 53% | GS&PD; wrist, MCP, PIP, flexor tendons (2-5 fingers), MTP2-5 & any other symptomatic joints | sonorheumatologist  (6y experience of MSUS) | ACR 1987 or ACR 2010; min 12 mos  RA 38, 37%; other confirmed dx 7, 7%; UA 22, 21%; no specific dx 36, 35% (referred back to GP) |
| **Sahbudin 2018 ^18^**  N = 107 | synovitis >=1 joint; <= 3 mos (RA 7 wks; IQR 5-9 wks. non-RA 5 wks; IQR 4-8 wks. resolving RA 5 wks; IQR 3-7 wks) | NR (incl in prior paper) | | Incl (43, 40%) | 60, 56%  RA; non-RA; res- olving  61 y; 39 y; 44 y§ | NR  NR | RA; non-RA; resolving  7 (IQR 3, 11); 2 (IQR 1, 6); 2 (IQR 1, 5)  12 (IQR 4, 15); 5 (IQR 2, 10); 5 (IQR 2, 7) | NR‡ | GS&PD; 19 bilateral joint sites, 16 bilateral tendon compartments (MCP 1-5, PIP 1-5 plus wrist, elbow, shoulder, knee, ankle) | rheumatologist | ACR 2010; min FU 18 mos  RA 46, 43%; other confirmed dx 37, 35% (incl 23/25 resolving); UA 24, 22% (incl 21/24 resolving) |
| **Salaffi 2010 ^19^**  N = 149 | synovitis >=1 joint wrist or finger & >=1 of: RF+, aCCP +, early morning stiffness >30 minutes or +ve MTPj squeeze test; <16 wks (mean 10.5 wks) | Excl | | N/A | 108, 72%  57y | NR  NR | NR  NR | 65, 44%‡ | GS&PD; Wrists, hands (MCP2-5, PIP2-5) | single experienced rheumatologist | ACR 1987; mean 12 mos (11 to 14 mos)  RA 62, 42%; other confirmed dx 18, 12%; UA 47, 32%; resolving 22, 15% |

ACR – American College of Rheumatology; anti-CCP – anti-cyclic citrullinated peptide/protein antibodies; BMO – bone marrow oedema; CI – confidence interval; d – days; DMARD – disease-modifying antirheumatic drugs; Dx – diagnosis; ERA - early rheumatoid arthritis (3-12 months duration); EULAR – European League Against Rheumatism; Excl – excluded; GS – grey scale; Incl – included; IQR – inter-quartile range; MCP – metacarpophalangeal; min – minimum; MRI – magnetic resonance imaging; MTP – metatarsophalangeal; N – number; N/A – not applicable; NR – not reported; OMERACT - Outcomes Measures in Rheumatoid Arthritis Clinical Trials; PD – power Doppler; PIP - proximal interphalangeal; RA – rheumatoid arthritis; RF – rheumatoid factor; T – Tesla; UA – unclassified arthritis; US – ultrasound; VERA - very early rheumatoid arthritis (<3 months duration); wks – weeks; mos – months; y – years.

* assume participants meeting ACR 2010 at baseline excluded as per UA definition in prior study ^20^

† participants meeting ACR 1987 at baseline were excluded by review team

‡ excluded at baseline if prior DMARD received

§ median age

|Methods section describes reference standard as fulfilment of 1987 ACR criteria or ongoing DMARD treatment, however Results section clearly describes accuracy estimates against reference standard of a clinical diagnosis classification of RA according to the predefined criteria

¶ PD also performed but data not presented as 2x2 and results not provided by author

**Reference list**

1.	Aoki T, Yamashita Y, Saito K, Tanaka Y, Korogi Y. Diagnosis of early-stage rheumatoid arthritis: usefulness of unenhanced and gadolinium-enhanced MR images at 3 T. *Clinical Imaging.* 2013;37(2):348-353.

2.	Boer AC, Burgers LE, Mangnus L, et al. Using a reference when defining an abnormal MRI reduces false-positive MRI results-a longitudinal study in two cohorts at risk for rheumatoid arthritis. *Rheumatology (Oxford).* 2017;56(10):1700-1706.

3.	Boeters DM, Boer AC, Van Der Helm-Van Mil AHM. Evaluation of the predictive accuracy of MRI-detected erosions in hand and foot joints in patients with undifferentiated arthritis. *Annals of the Rheumatic Diseases.* 2019;78(1):144–146.

4.	Duer A, Ostergaard M, Horslev-Petersen K, Vallo J. Magnetic resonance imaging and bone scintigraphy in the differential diagnosis of unclassified arthritis. *Annals of the Rheumatic Diseases.* 2008;67(1):48-51.

5.	Duer-Jensen A, Horslev-Petersen K, Hetland ML, et al. Bone edema on magnetic resonance imaging is an independent predictor of rheumatoid arthritis development in patients with early undifferentiated arthritis. *Arthritis & Rheumatism.* 2011;63(8):2192-2202.

6.	Ji L, Li G, Xu Y, Zhou W, Zhang Z. Early prediction of rheumatoid arthritis by magnetic resonance imaging in the absence of anti-cyclic citrullinated peptide antibodies and radiographic erosions in undifferentiated inflammatory arthritis patients: a prospective study. *International Journal of Rheumatic Diseases.* 2015;18(8):859-865.

7.	Klarlund M, Ostergaard M, Jensen KE, Madsen JL, Skjodt H, Lorenzen I. Magnetic resonance imaging, radiography, and scintigraphy of the finger joints: One year follow up of patients with early arthritis. *Annals of the Rheumatic Diseases.* 2000;59(7):521-528.

8.	Lei X, Li H, Zhan Y, Qu J. Predict rheumatoid arthritis conversion from undifferentiated arthritis with dynamic contrast-enhanced MRI and laboratory indexes. *Clinical & Experimental Rheumatology.* 2018;36(4):552-558.

9.	Li R, Liu X, Ye H, et al. Magnetic resonance imaging in early rheumatoid arthritis: a multicenter, prospective study. *Clinical Rheumatology.* 2016;35(2):303-308.

10.	Mori G, Tokunaga D, Takahashi KA, et al. Maximum intensity projection as a tool to diagnose early rheumatoid arthritis. *Modern Rheumatology.* 2008;18(3):247-251.

11.	Narvaez J, Sirvent E, Narvaez JA, et al. Usefulness of magnetic resonance imaging of the hand versus anticyclic citrullinated peptide antibody testing to confirm the diagnosis of clinically suspected early rheumatoid arthritis in the absence of rheumatoid factor and radiographic erosions. *Seminars in Arthritis & Rheumatism.* 2008;38(2):101-109.

12.	Solau-Gervais E, Legrand JL, Cortet B, Duquesnoy B, Flipo RM. Magnetic resonance imaging of the hand for the diagnosis of rheumatoid arthritis in the absence of anti-cyclic citrullinated peptide antibodies: a prospective study. *Journal of Rheumatology.* 2006;33(9):1760-1765.

13.	Tamai M, Kawakami A, Uetani M, et al. A prediction rule for disease outcome in patients with undifferentiated arthritis using magnetic resonance imaging of the wrists and finger joints and serologic autoantibodies. *Arthritis & Rheumatism.* 2009;61(6):772-778.

14.	Horton SC, Tan AL, Wakefield RJ, Freeston JE, Buch MH, Emery P. Ultrasound-detectable grey scale synovitis predicts future fulfilment of the 2010 ACR/EULAR RA classification criteria in patients with new-onset undifferentiated arthritis. *RMD Open.* 2017;3(1):e000394.

15.	Ji L, Deng X, Geng Y, Song Z, Zhang Z. The additional benefit of ultrasonography to 2010 ACR/EULAR classification criteria when diagnosing rheumatoid arthritis in the absence of anti-cyclic citrullinated peptide antibodies. *Clinical Rheumatology.* 2017;36(2):261-267.

16.	Ponikowska M, Swierkot J, Nowak B. The importance of ultrasound examination in early arthritis. *Reumatologia.* 2018;56(6):354-361.

17.	Rezaei H, Torp-Pedersen S, af Klint E, et al. Diagnostic utility of musculoskeletal ultrasound in patients with suspected arthritis--a probabilistic approach. *Arthritis Research & Therapy.* 2014;16(5):448.

18.	Sahbudin I, Pickup L, Nightingale P, et al. The role of ultrasound-defined tenosynovitis and synovitis in the prediction of rheumatoid arthritis development. *Rheumatology (Oxford).* 2018;57(7):1243-1252.

19.	Salaffi F, Ciapetti A, Gasparini S, Carotti M, Filippucci E, Grassi W. A clinical prediction rule combining routine assessment and power Doppler ultrasonography for predicting progression to rheumatoid arthritis from early-onset undifferentiated arthritis. *Clinical & Experimental Rheumatology.* 2010;28(5):686-694.

20.	Nieuwenhuis WP, Mangnus L, van Steenbergen HW, et al. Older age is associated with more MRI-detected inflammation in hand and foot joints. *Rheumatology (Oxford).* 2016;55(12):2212-2219.

### Supplementary Figure 1 Additional details of QUADAS-2 assessment results

a. **Risk of bias and applicability concerns graph for studies evaluating MRI**

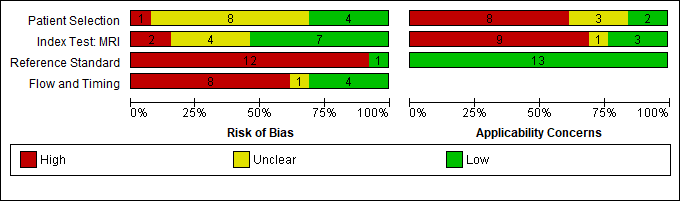

b. **Risk of bias and applicability concerns graph for studies evaluating ultrasound (US)**

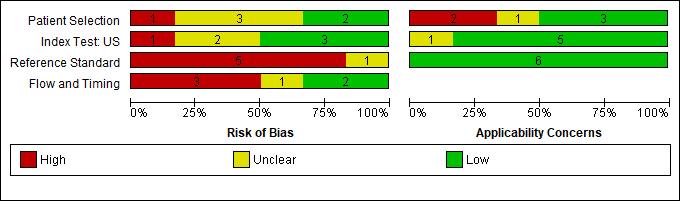

c. **Risk of bias and applicability concerns summary: review authors' judgements about each domain for each included study**

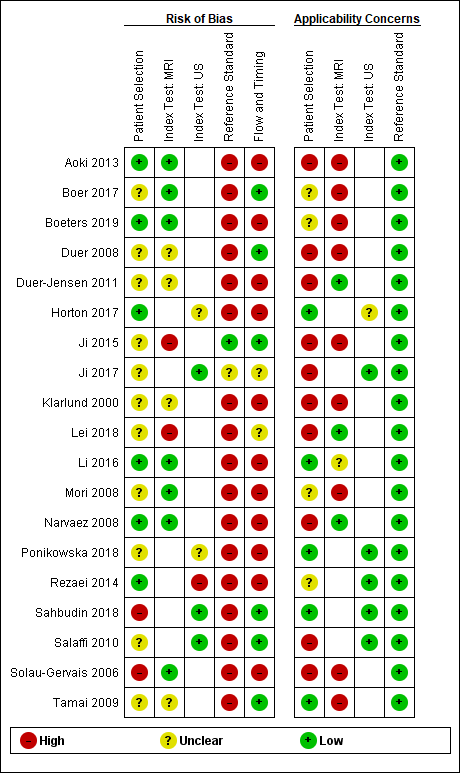

### Supplementary Figure 2 Forest plots of sensitivities and specificities from studies evaluating individual MRI characteristics at specific joints against ACR 1987 reference standard

a. **MCP joint**

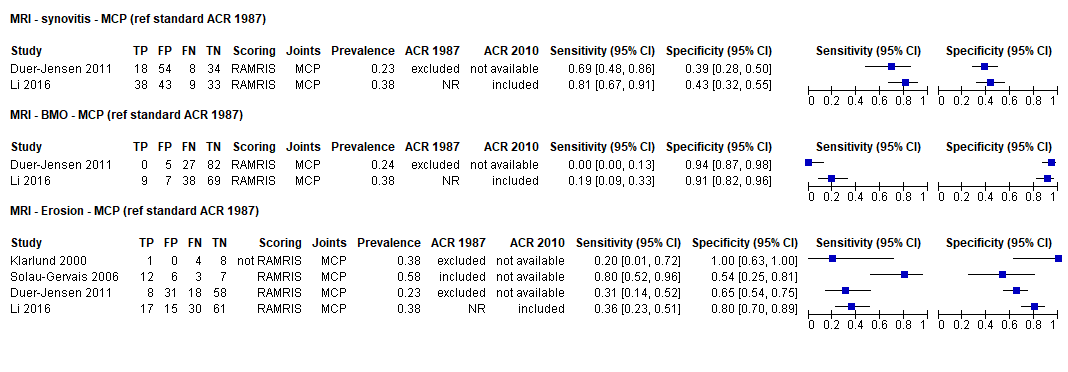

b. **PIP joint**

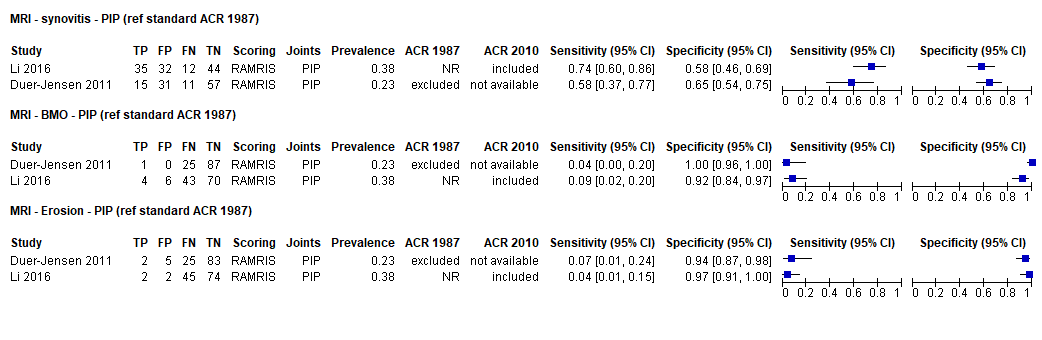

c. **Wrist**

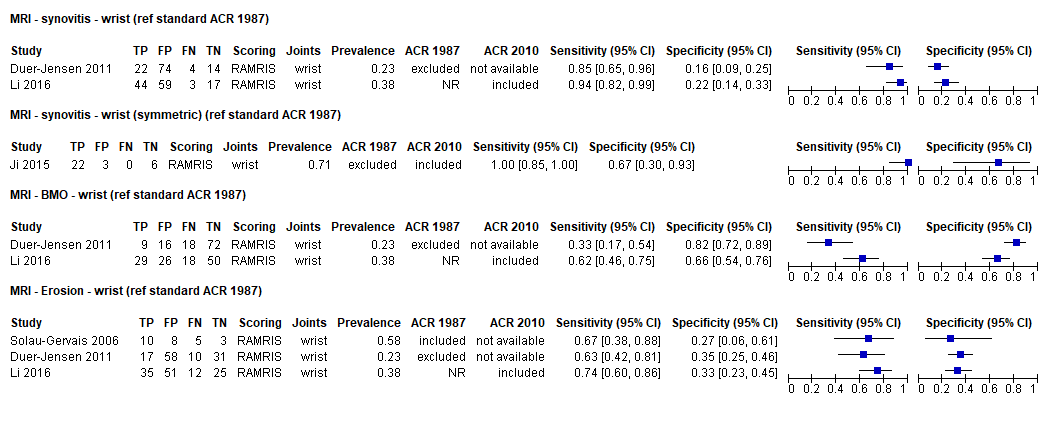

d. **MTP joint**

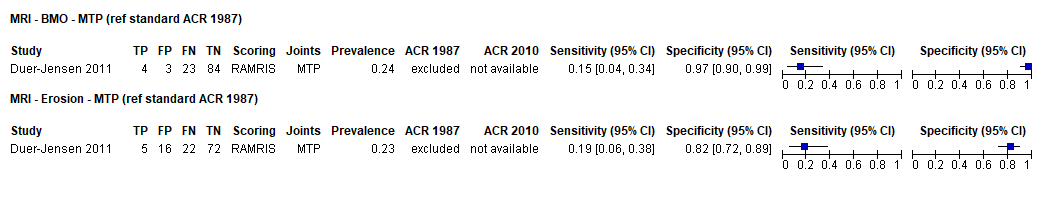

### Supplementary Figure 3 Forest plots of sensitivities and specificities from studies evaluating the accuracy of other MRI thresholds against ACR 1987 reference standard

a. **Any inflammation (various joint areas)**

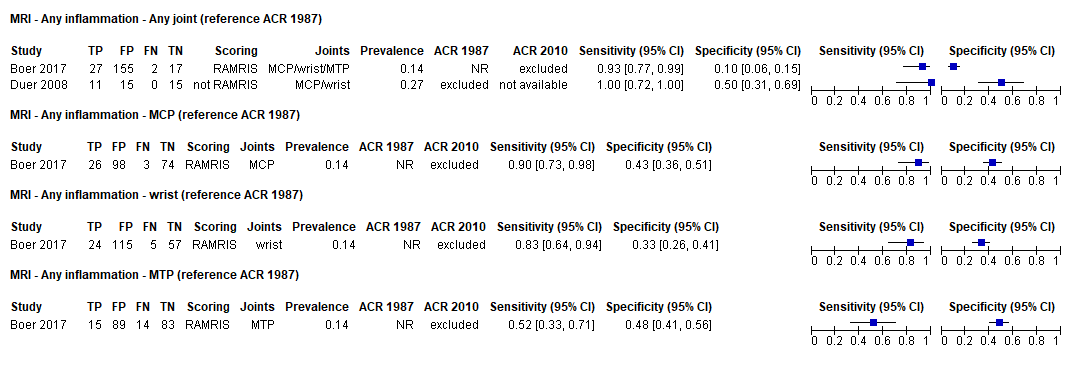

b. **Miscellaneous thresholds (ACR 1987 reference standard)**

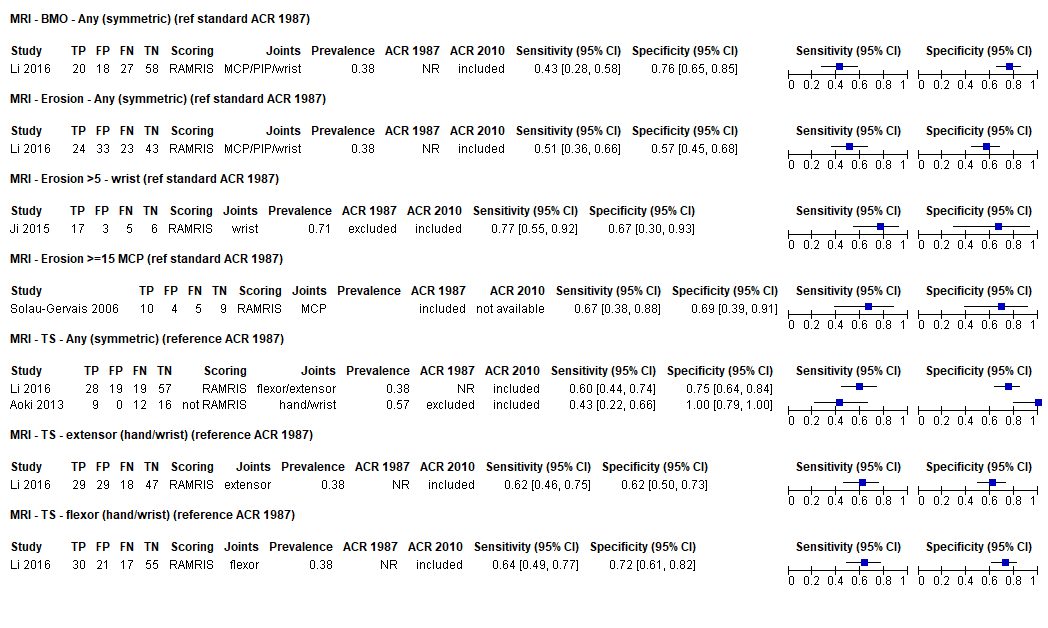

c. **Miscellaneous thresholds (mixed reference standards)**

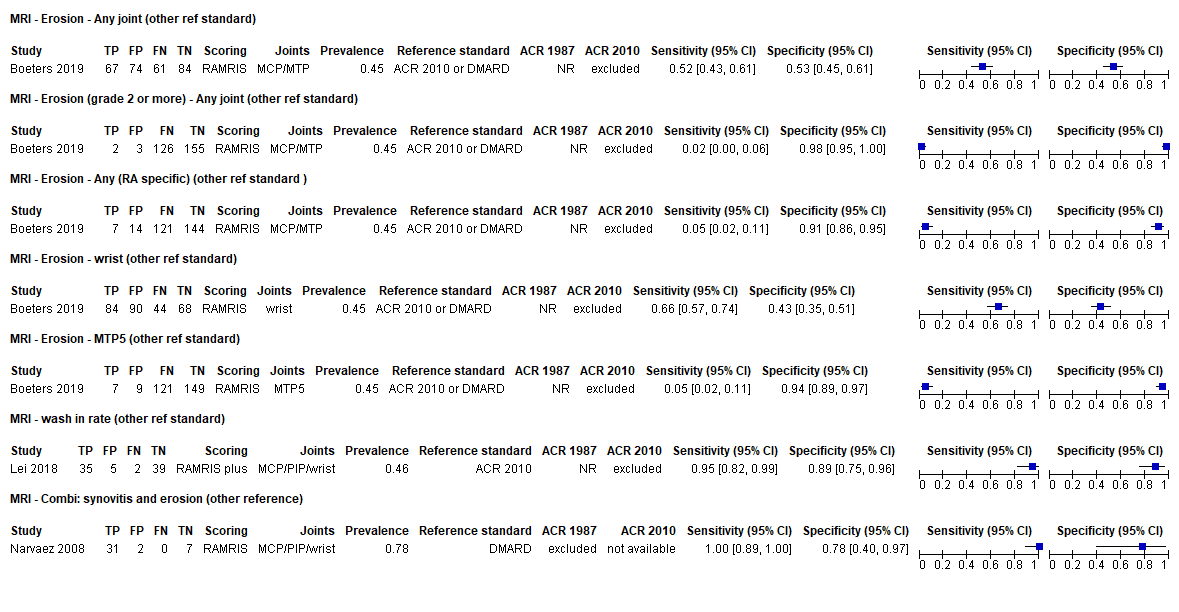

### Supplementary Figure 4 Forest plot of sensitivities and specificities from studies evaluating ultrasound (results across all joint areas)

a. Additional data for GS ultrasound synovitis or PD ultrasound synovitis alone

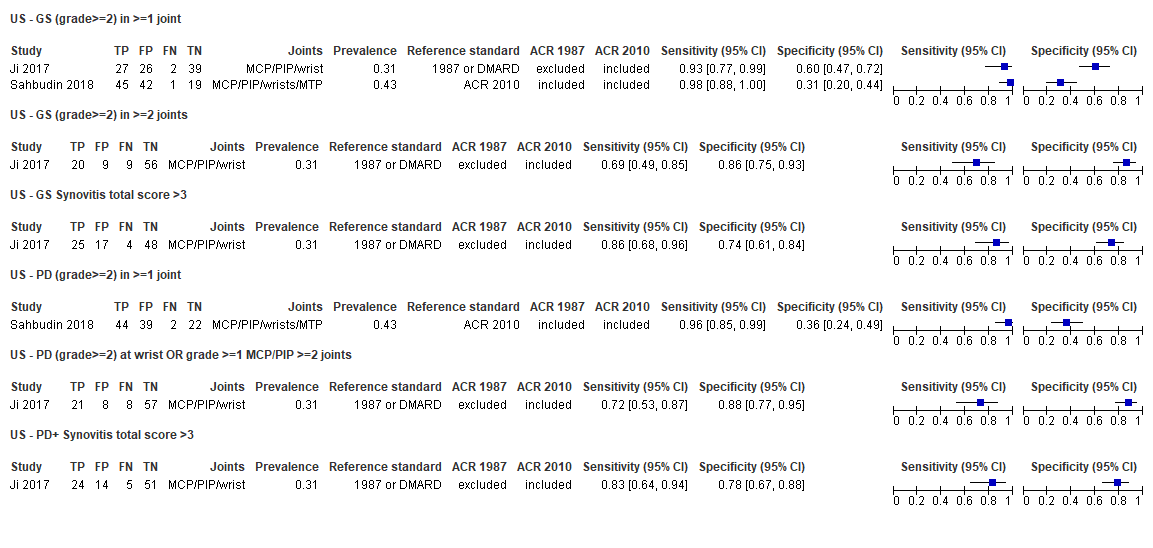

* All ultrasound evaluations included both grey scale (GS) synovitis and power dower (PD) activity. Synovitis was scored using similar semiquantitative grading scales.^24 48-50^

Two studies^53 57^ used OMERACT definition of synovitis^45 46^ and none used the EULAR-OMERACT combined scoring system for grading synovitis in RA^3-5^ .

b. Additional data for combinations of GS and PD ultrasound synovitis (positive in >=1 joint, or positive in multiple joints)

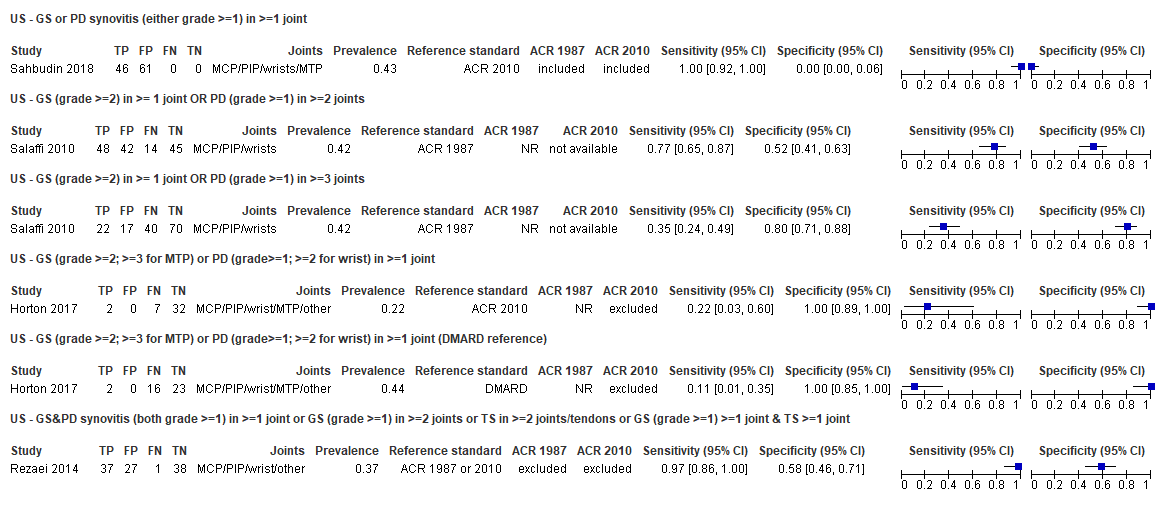

* All ultrasound evaluations included both grey scale (GS) synovitis and power dower (PD) activity. Synovitis was scored using similar semiquantitative grading scales.^24 48-50^

Two studies^53 57^ used OMERACT definition of synovitis^45 46^ and none used the EULAR-OMERACT combined scoring system for grading synovitis in RA^3-5^ .

c. Data for GS or PD tenosynovitis:

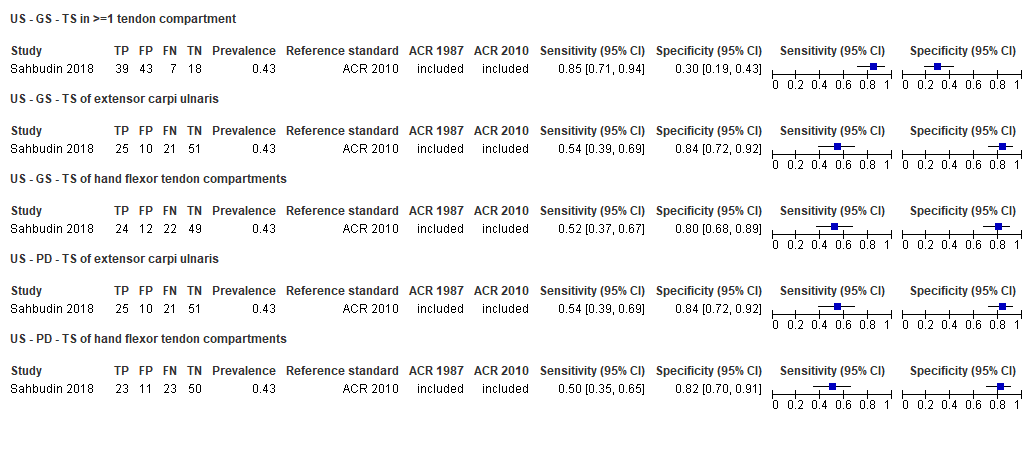

### Supplementary Figure 5 Forest plot of sensitivities and specificities from Sahbudin 2018 (results for individual joints and tendons)

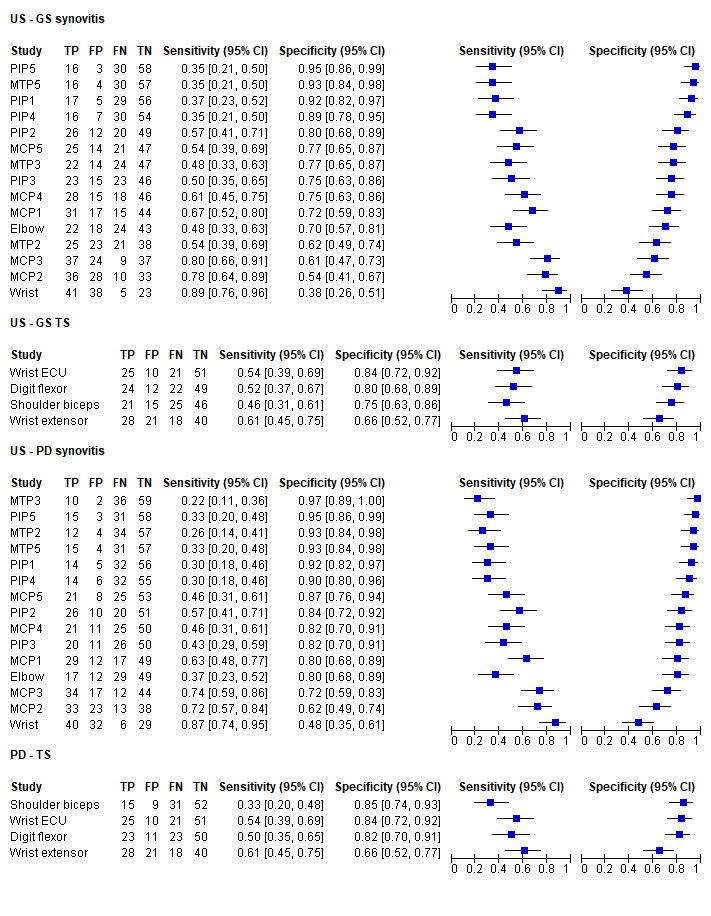

### Appendix 1. Early RA systematic review search strategies

**Database: Ovid MEDLINE(R) and In-Process & Other Non-Indexed Citations <1946 to May 01, 2019**

**Search date: 2^nd^ May 2019**

1	*Rheumatic diseases/pa, dg (640)

2	ARTHRITIS, RHEUMATOID/di, dg, cl, im [Diagnosis, Diagnostic Imaging, Classification] (31677)

3	((recent* or early or onset or suspect*or untreat* or undifferentiated or unclassified or inflammatory or asymptomatic or symptomatic) adj2 arthritis).ti,ab. (10178)

4	(((rheumatoid adj2 arthritis) or RA) adj2 (recent* or early or onset or suspect* or untreat*)).mp. [mp=title, abstract, original title, name of substance word, subject heading word, floating sub-heading word, keyword heading word, organism supplementary concept word, protocol supplementary concept word, rare disease supplementary concept word, unique identifier, synonyms] (5220)

5	polyarthrit*.ti,ab. (8852)

6	exp SYNOVITIS/ (7964)

7	synov*.ti,ab. and arthrit*.mp. (24376)

8	(arthralgia or myalgia).ti,ab. and arthrit*.mp. (1809)

9	((pain* or stiff*) adj3 joint*).ti,ab. and arthrit*.mp. (3329)

10	((pain* or stiff*) adj3 muscle*).ti,ab. and arthrit*.mp. (209)

11	AUTOANTIBODIES/ (64551)

12	Rheumatoid Factor/ (8659)

13	exp Anti-Citrullinated Protein Antibodies/ (131)

14	((autoantibody or autoantibodies) adj3 (test* or assay*)).ti,ab. (1740)

15	rheumatoid factor*.ti,ab. (10570)

16	(anti adj (CCP or CarP or carbamylate* or acetylate* or cyclic or mcv)).ti,ab. (2207)

17	(antibod* adj4 (CCP or filaggrin or antifilaggrin or anticyclic or citrull* or anticitrull or vimentin)).ti,ab. (4036)

18	11 or 12 or 13 or 14 or 15 or 16 or 17 (79387)

19	magnetic resonance imaging/ or echo-planar imaging/ (375974)

20	(MRI or "magnetic resonance imag*" or "magnetic resonance probe").ti,ab. (327240)

21	ULTRASONOGRAPHY, DOPPLER/ or ULTRASONOGRAPHY, DOPPLER, PULSED/ or ULTRASONOGRAPHY/ or ULTRASONOGRAPHY, DOPPLER, DUPLEX/ or ULTRASONOGRAPHY, DOPPLER, COLOR/ (207027)

22	(ultrasonog* or ultrasound).ti,ab. (295876)

23	exp Tomography, Emission-Computed/ (104437)

24	Positron Emission Tomography Computed Tomography/ (5968)

25	Single Photon Emission Computed Tomography Computed Tomography/ (630)

26	Optical Imaging/ (7396)

27	Lymphoscintigraphy/ (895)

28	exp Tomography, Emission-Computed/ (104437)

29	(single photon emission computed tomography or SPECT).ti,ab. (29680)

30	(positron emission tomography or PET).ti,ab. (100386)

31	(diffuse optical* adj2 transmiss*).ti,ab. (3)

32	(DOI or NIROT or FDOT).ti,ab. (14816)

33	((diffuse optical* or NIR or Near-infrared or Near infrared or fluorescen* or indocyanine green or ICG) adj2 (imag* or probe or spectroscop*)).ti,ab. (63984)

34	(Xiralite or Handscan).mp. (7)

35	scintigraph*.ti,ab. (45034)

36	(photoacoustic imag* or photoacoustic tomography).ti,ab. (1729)

37	(Intelligent adj3 probe).ti,ab. (1)

38	19 or 20 or 21 or 22 or 23 or 24 or 25 or 26 or 27 or 28 or 29 or 30 or 31 or 32 or 33 or 34 or 35 or 36 or 37 (1085719)

39	(predict* or clinical or outcome* or risk*).mp. (7317369)

40	(rule* or algorithm* or model*).ti,ab. (2818165)

41	39 and 40 (1079503)

42	1 or 2 or 3 or 4 or 5 or 6 or 7 or 8 or 9 or 10 [mp=title, abstract, original title, name of substance word, subject heading word, floating sub-heading word, keyword heading word, organism supplementary concept word, protocol supplementary concept word, rare disease supplementary concept word, unique identifier, synonyms] (73193)

43	18 or 38 or 41 (2181983)

44	42 and 43 (17377)

45	Animals/ (6393978)

46	Humans/ (17695575)

47	45 not (45 and 46) (4541425)

48	44 not 47 (16190)

49	limit 48 to yr="1987-Current" (13662)

**Database: Embase <1974 to 2019 May 01>**

**Search date: May 2^nd^ 2019**

1	*Rheumatic disease/di (1755)

2	*rheumatoid arthritis/di (9260)

3	((recent* or early or onset or suspect*or untreat* or undifferentiated or unclassified or inflammatory or asymptomatic or symptomatic) adj2 arthritis).ti,ab. (18893)

4	(((rheumatoid adj2 arthritis) or RA) adj2 (recent* or early or onset or suspect* or untreat*)).ti,ab. [mp=title, abstract, heading word, drug trade name, original title, device manufacturer, drug manufacturer, device trade name, keyword, floating subheading word, candidate term word] (9993)

5	polyarthrit*.ti,ab. (8938)

6	*SYNOVITIS/ (4661)

7	(synov* and arthrit*).ti,ab. (32146)

8	((arthralgia or myalgia) and arthrit*).ti,ab. (3355)

9	(((pain* or stiff*) adj3 joint*) and arthrit*).ti,ab. (5794)

10	(((pain* or stiff*) adj3 muscle*) and arthrit*).ti,ab. (402)

11	*AUTOANTIBODY/ (28992)

12	*Rheumatoid Factor/ (3991)

13	exp cyclic citrullinated peptide antibody/ (5122)

14	((autoantibody or autoantibodies) adj3 (test* or assay*)).ti,ab. (2751)

15	rheumatoid factor*.ti,ab. (16072)

16	(anti adj (CCP or CarP or carbamylate* or acetylate* or cyclic or mcv)).ti,ab. (5632)

17	(antibod* adj4 (CCP or filaggrin or antifilaggrin or anticyclic or citrull* or anticitrull or vimentin)).ti,ab. (7535)

18	11 or 12 or 13 or 14 or 15 or 16 or 17 (53612)

19	nuclear magnetic resonance imaging/ or echo planar imaging/ (721698)

20	(MRI or "magnetic resonance imag*" or "magnetic resonance probe").ti,ab. (515899)

21	DOPPLER ultrasonography/ or PULSED DOPPLER ULTRASONOGRAPHY/ or ECHOGRAPHY/ or ULTRASONOGRAPHY, DOPPLER, DUPLEX/ or COLOR DOPPLER FLOWMETRY/ (305617)

22	(ultrasonog* or ultrasound).ti,ab. (450006)

23	exp computer assisted emission tomography/ (175878)

24	Positron Emission Tomography Computed Tomography/ (18959)

25	Single Photon Emission Computed Tomography-Computed Tomography/ (2514)

26	fluorescence Imaging/ (19617)

27	Lymphoscintigraphy/ (5493)

28	exp computer assisted emission tomography/ (175878)

29	(single photon emission computed tomography or SPECT).ti,ab. (48577)

30	(positron emission tomography or PET).ti,ab. (171651)

31	(diffuse optical* adj2 transmiss*).ti,ab. (4)

32	(DOI or NIROT or FDOT).ti,ab. (39322)

33	((diffuse optical* or NIR or Near-infrared or Near infrared or fluorescen* or indocyanine green or ICG) adj2 (imag* or probe or spectroscop*)).ti,ab. (75744)

34	(Xiralite or Handscan).ti,ab. (33)

35	scintigraph*.ti,ab. (60963)

36	(photoacoustic imag* or photoacoustic tomography).ti,ab. (1977)

37	(Intelligent adj3 probe).ti,ab. (3)

38	19 or 20 or 21 or 22 or 23 or 24 or 25 or 26 or 27 or 28 or 29 or 30 or 31 or 32 or 33 or 34 or 35 or 36 or 37 (1708628)

39	(predict* or clinical or outcome* or risk*).ti,ab. (8581344)

40	(rule* or algorithm* or model*).ti,ab. (3623222)

41	39 and 40 (1470505)

42	1 or 2 or 3 or 4 or 5 or 6 or 7 or 8 or 9 or 10 [mp=title, abstract, heading word, drug trade name, original title, device manufacturer, drug manufacturer, device trade name, keyword, floating subheading word, candidate term word] (75258)

43	18 or 38 or 41 (3132129)

44	42 and 43 (21171)

45	Animals/ (1212281)

46	Humans/ (11290293)

47	45 not (45 and 46) (947683)

48	44 not 47 (21076)

49	limit 48 to yr="1987-Current" (20438)

**Database: Web of Science: Biosis Citation Index**

**Search date: 7^th^ May 2019**

#1 TS=Animal* (5,265,010)

#2 TS=Human* (3,900,849)

#3 #1 NOT (#1 and #2) (1,662,450)

#4 TS="Rheumatic disease*" (5,045)

#5 TS=" Arthritis, Rheumatoid" (38,444)

#6 TS="Rheumatoid arthritis" (42,813)

#7 TS=(Synov*) (14,413)

#8 TS=(Polyarthrit*) (1,245)

#9 #8 OR #7 OR #6 OR #5 OR #4 (55,358)

#10 TS=("magnetic resonance probe”) (14)

#11 TS=(ultrasonog* or ultrasound) (84,138)

#12 TS=("single photon emission computed tomography" or SPECT) (8,891)

#13 TS=("positron emission tomography" or PET) (51,455)

#14 TS= ("diffuse optical* transmiss*") (2)

#15 TS= (DOI or NIROT or FDOT) (47,618)

#16 TS=(Xiralite or Handscan) (15)

#17 TS=(scintigraph*) (5,032)

#18 TS= ("photoacoustic imag*" or "photoacoustic tomography") (998)

#19 TS=(“Intelligent probe”) (2)

#20 TS=(“magnetic resonance imag*”)(123,474)

#21 TS=("diffuse optical*" or "NIR" or "Near-infrared" or "Near infrared" or "fluorescen*" or "indocyanine green" or "ICG") (223,520)

#22 #21 OR #20 OR #19 OR #18 OR #17 OR #16 OR #15 OR #14 OR #13 OR #12 OR #11 OR #10 (512,751)

#23 #22 AND #9 (5,687)

#24 #23 NOT #3 (5,272)

#25 TS=("predict*" or "clinical" or "outcome*" or "risk*") (3,747,085)

#26 TS=("rule*" or "algorithm" or "model*") (1,309,120)

#27 #26 AND #25 (719,356)

#28 TS= (Autoantibody) (16,080)

#29 TS=("Rheumatoid Factor") (4,062)

#30 TS=("cyclic citrullinated peptide antibody") (456)

#31 TS=(("autoantibod*") Near/3 ("test*" or "assay*")) (632)

#32 TS=("rheumatoid factor*") (4,124)

#33 TS=("anti" NEAR ("CCP" or "CarP" or "carbamylate" or "acetylate*" or "cyclic" or "mcv"))(2,728)

#34 TS=(antibod* NEAR/4 ("CCP" or "filaggrin" or "antifilaggrin" or "anticyclic" or "citrull*" or "anticitrull" or "vimentin")) (2,985)

#35 #34 OR #33 OR #32 OR #31 OR #30 OR #29 OR #28 (22,170)

#36 #35 OR #27 OR #22 (1,193,351)

#37 #36 AND #9 (16,488)

#38 #37 not #3 (15,180)

### Appendix 2. QUALITY ASSESSMENT: RA DIAGNOSIS

The QUADAS-2 checklist (Whiting 2011) was tailored to the review topic as follow:.

| **Domain** | **Response options** |
| --- | --- |
| **Participant selection - Risk of bias** |  |
| **1. Was a consecutive or random sample of participants or images enrolled?** | Yes - if paper states consecutive or random, or if all participants meeting explicit study eligibility criteria within a specified time frame were included  No – if paper describes other method of sampling Unclear – if participant sampling not described |
| **2. Did the study avoid inappropriate exclusions** | Yes - if no inappropriate exclusion criteria were applied  No – if participant exclusion criteria were applied that might bias results or affect test accuracy in some way  Unclear – if not clearly reported |
| **Could the selection of participants have introduced bias?** | Low risk if all responses are Yes High risk if one or more responses are No Unclear risk if one or more responses are Unclear , and no responses are No |
| **Participant selection - Concerns about applicability** |  |
| **Are the included patients and chosen study setting appropriate to answer the review question, i.e. are the study results generalisable?** | Low concern – if study eligiblity criteria met review defintion for UA and data can be extracted for participants with unclassified arthritis on study entry, and median symptom duration was < 12 months at baseline or >=50% of recruited participants have symptom duration < 1 year High concern - if study eligiblity criteria did not meet review defintion for UA (e.g. symmetric synovitis required or invovlement of more than 2 joints required) or median symptom duration was > 12 months at baseline or >=50% of recruited participants have symptom duration < 1 year Unclear concern – if insufficient details are provided |
| **Index test - Risk of bias** |  |
| **1. Was the index test or testing strategy result interpreted without knowledge of the results of the reference standard (final patient diagnosis)?** | Always Yes unless images acquired at baseline were re-interpreted for purposes of the study, in which case blinding must be clearly stated to score Yes |
| **2. Was the diagnostic threshold at which the index test was considered positive pre-specified?** | Yes - if threshold was pre-specified (i.e. prior to analysing study results), such that study results were not data driven. For imaging tests, reported use of RAMRIS or OMERACT scoring will be considered sufficient, even if the specific grade required for a positive test result is not reported No - if threshold was not pre-specified but was selected after analysis of results, usually to maximise sensitivity and/or specificity, or multiple thresholds were tested to identify the optimal threshold Unclear - if not possible to tell whether or not diagnostic threshold was pre-specified |
| **Could the conduct or interpretation of the index test have introduced bias?** | Low risk if all responses are Yes High risk if one or more responses are No Unclear risk if one or more responses are Unclear , and no responses are No |
| **Index - Concerns about applicability** |  |
| **Are there concerns that the index test, its conduct, or interpretation differ from the review question?** | High concern present if test was interpreted by more than one reader (consensus or mean result used) or if intepretation was blinded to other clinical information |
| **Reference standard - Risk of bias** |  |
| **1. Is the reference standard likely to correctly classify the target condition?** | Yes – if the ACR criteria were clearly assessed for all included participants or if the presence of RA was defined by initiation of DMARD therapy.  No – if any disease positive participant had a final diagnosis of RA made by expert diagnosis or consensus opinion OR if any disease positive participant did not fully meet the ACR criteria for RA OR if the ACR criteria were not explicitly assessed for the RA absent group  Unclear – if the method of final diagnosis was not reported for any disease positive participant |
| **2. Were the reference standard results interpreted without knowledge of the results of the index test?** | Yes – if the final subjective assessment of the presence of RA (e.g. swollen joint counts) was made blinded to the original index test results No – if the final subjective assessment of the presence of RA (e.g. swollen joint counts) was made in the knowledge of the original index test result (if blinding is not specifically stated (e.g. joint assessment by a research nurse blinded to assay results) then we should assume that baseline index test results are known and respond No to this item) Unclear – if blinded reference test interpretation was not clearly reported |
| **Could the reference standard, its conduct, or its interpretation have introduced bias?** | Low risk if all responses are Yes High risk if one or more responses are No Unclear risk if one or more responses are Unclear , and no responses are No |
| **Reference standard - Concerns about applicability** |  |
| **Are there concerns that the target condition as defined by the reference standard does not match the question?** |  |
| **Flow and timing** |  |
| **1. Was there an appropriate interval between index test and reference standard?** | Yes – if study specifes a minimum of at least 1 year of follow-up (even if some lost to FU) No – if study reports <1 year follow-up  Unclear – if study does not report length of clinical follow-up |
| **2. Did all included participants receive an eligible reference standard?** | Yes – if all participants underwent an eligible reference standard  No – – if some participants did not receive an eligible reference standard  Unclear – if not clearly reported |
| **3. Did all participants receive the same reference standard?** | Yes – if only one reference standard was used, e.g. only ACR 2010, or only DMARD initiation  No – if more than one reference standard was used Unclear – if not clearly reported |
| **4. Were all participants included in the analysis?** | Yes – if all participants who were recruited into the study (and who meet our eligibility criteria) were included in the analysis  No – if some participants who were recruited into the study were later excluded from the analysis (e.g. loss to follow-up) Unclear– if not clearly reported |
| **Could the patient flow have introduced bias?** | Low risk if all responses are Yes High risk if one or more responses are No Unclear risk if one or more responses are Unclear , and no responses are No |
